# Relationships between depression, anxiety, and motivation in the real-world: Effects of physical activity and screentime

**DOI:** 10.1101/2024.08.06.24311477

**Authors:** J. Beltrán, Y. Jacob, M. Mehta, T. Hossain, A. Adams, S. Fontaine, J. Torous, C. McDonough, M. Johnson, A. Delgado, J. W. Murrough, L. S. Morris

**Author notes:** Correspondence: Laurel S. Morris Department of Psychiatry, Icahn School of Medicine at Mount Sinai, One Gustave L. Levy Pl., New York, NY.

## Abstract

**Background:** Mood and anxiety disorders are highly prevalent and comorbid worldwide, with variability in symptom severity that fluctuates over time. Digital phenotyping, a growing field that aims to characterize clinical, cognitive and behavioral features via personal digital devices, enables continuous quantification of symptom severity in the real world, and in real-time.

**Methods:** In this study, N=114 individuals with a mood or anxiety disorder (MA) or healthy controls (HC) were enrolled and completed 30-days of ecological momentary assessments (EMA) of symptom severity. Novel real-world measures of anxiety, distress and depression were developed based on the established Mood and Anxiety Symptom Questionnaire (MASQ). The full MASQ was also completed in the laboratory (in-lab). Additional EMA measures related to extrinsic and intrinsic motivation, and passive activity data were also collected over the same 30-days. Mixed-effects models adjusting for time and individual tested the association between real-world symptom severity EMA and the corresponding full MASQ sub-scores. A graph theory neural network model (D_EP_NA) was applied to all data to estimate symptom interactions.

**Results:** There was overall good adherence over 30-days (MA=69.5%, HC=71.2% completion), with no group difference (t_(58)_=0.874, p=0.386). Real-world measures of anxiety/distress/depression were associated with their corresponding MASQ measure within the MA group (t’s > 2.33, p’s < 0.024). Physical activity (steps) was negatively associated with real-world distress and depression (IRRs > 0.93, p’s ≤ 0.05). Both intrinsic and extrinsic motivation were negatively associated with real-world distress/depression (IRR’s > 0.82, p’s < 0.001). D_EP_NA revealed that both extrinsic and intrinsic motivation significantly influenced other symptom severity measures to a greater extent in the MA group compared to the HC group (extrinsic/intrinsic motivation: t_(46)_ = 2.62, p < 0.02, q FDR < 0.05, Cohen’s *d* = 0.76; t_(46)_ = 2.69, p < 0.01, q FDR < 0.05, Cohen’s *d* = 0.78 respectively), and that intrinsic motivation significantly influenced steps (t_(46)_ = 3.24, p < 0.003, q FDR < 0.05, Cohen’s *d* = 0.94).

**Conclusions:** Novel real-world measures of anxiety, distress and depression significantly related to their corresponding established in-lab measures of these symptom domains in individuals with mood and anxiety disorders. Novel, exploratory measures of extrinsic and intrinsic motivation also significantly related to real-world mood and anxiety symptoms and had the greatest influencing degree on patients’ overall symptom profile. This suggests that measures of cognitive constructs related to drive and activity may be useful in characterizing phenotypes in the real-world.

## Introduction

Mood and anxiety disorders are highly prevalent and comorbid in the United States (Hasin et al., 2018; Saha et al., 2021). Mood disorders such as Major Depressive Disorder (MDD) and anxiety disorders such as Generalized Anxiety Disorder (GAD) are characterized by a defined set of psychiatric symptoms, but both categorical diagnoses share substantial overlap in their clinical presentation. For example, both present with executive function deficits, sleep disturbances, fatigue, maladaptive arousal, and psychomotor abnormalities (Liu et al., 2018). This overlap, co-occurrence (Kessler et al., 2015), and mutual exacerbation (Liu et al., 2018) can complicate specificity of diagnoses and measurement of treatment outcomes. Measurement of symptom severity rather than categorical diagnoses can thus be useful in characterizing these overlapping disorder domains.

Current tools for measuring and diagnosing mood and anxiety disorders include clinical interview, often via the Diagnostic and Statistical Manual of Mental Disorders (DSM), which facilitates categorical diagnosis of disorders. The self-reported Mood and Anxiety Symptom Questionnaire (MASQ) (Watson et al., 1995) can also measure symptom severity for separate domains of anxious arousal, general distress, and anhedonic depression on a dimensional continuum. While this assessment can discriminate anxiety and depressive symptom severity within psychiatric patient populations (Watson et al., 1995), much like many other self-report questionnaires, the MASQ requires that a patient recall a week’s worth of experiences and distill complex emotional and mental states into a single number. This recall bias may therefore lead to an inaccurate representation of symptom severity that has been averaged over a long period over time and potentially distorted by any underlying cognitive dysfunction, which is highly prevalent in mood and anxiety disorders (Balderston et al., 2017; Gadassi Polack et al., 2020; Khdour et al., 2016; LeMoult and Gotlib, 2019; Onnela, 2021). Determining diagnosis and symptom severity in this way overlooks the intricacies and nuances of symptom profiles that can fluctuate on a day-to-day basis within patients (Nemesure et al., 2022; van Eeden et al., 2019).

Digital phenotyping – i.e. momentary assessment of symptom profiles in the real-world through personal digital devices – can counteract some of these limitations in symptom profiling, by assessing symptom severity across multiple timepoints, with low patient burden (Torous et al., 2016). Approximately 90% of Americans own a smartphone (“Mobile Fact Sheet,” 2024) with a projected increase to over 5 billion people owning smartphones globally by 2030 (Poushter, 2016; Reinertsen and Clifford, 2018). The ubiquity of smartphones enables novel quantification of patients’ behaviors and symptoms in real life and real-time through digital phenotyping. Since mood and anxiety symptom severity can fluctuate over time, digital phenotyping tools may detect vulnerability towards mood and anxiety disorders before they become taxing on an individual’s life, and indicate behavioral markers that predict symptom changes (Choi et al., 2024). Digital phenotyping tools may also highlight how different symptoms interact with one another (Ebrahimi et al., 2024), and how underlying cognitive changes, such as changes in motivation, might impact those relationships (Fervaha et al., 2016). Maladaptive changes in motivation are consistently observed in individuals with mood and anxiety disorders (Bi et al., 2022; Charpentier et al., 2017; Treadway et al., 2012; Westra et al., 2009), and recent evidence suggests that both intrinsic motivation (actions driven by ‘internal drivers’) and extrinsic motivation (actions driven by tangible external stimuli or outcomes) may be differentially impaired (Morris et al., 2022). While motivational deficits are commonly observed within psychiatric populations, understanding how different kinds of motivation may differentially impact symptom severity in the real-world remains limited.

Over the past decade, research on digital phenotyping has accelerated (Choi et al., 2024) by providing active data (captured via ecological momentary assessments (EMAs) (Shiffman et al., 2008) and passive data from sensors including accelerometer, GPS, and screen usage that can provide greater ecological validity and minimize patient recall bias. The utility of digital phenotyping has been exemplified across many psychiatric conditions (Bufano et al., 2023). Wearable sensors can provide motion data that helps infer the risk of anxiety disorders such as GAD (Jacobson and Feng, 2022), although wearable devices can be costly and could exclude certain populations. Motion data from phone sensors alone also holds value and may be able to inform predictions of periods of increased anxiety (Cohen et al., 2024; Nguyen et al., 2022). Taken together these studies suggest that digital phenotyping, and in particular passively collected sensor data, can be used to probe psychiatric symptoms and serve as a powerful tool for enabling early detection of behaviors that may inform risk prediction.

The current study presents a new set of single-item, self-reported, real-world measures of anxiety, distress, and depression captured via a research-based open-source smartphone application (*mindLAMP*) (Vaidyam et al., 2022) in individuals with mood and anxiety (MA) disorders and healthy controls (HC). Gold-standard clinical assessments of mood and anxiety symptom severity (MASQ) were also captured in the laboratory (in-lab) in order to test whether real-world measures of Anxious Arousal (anxiety), General Distress (distress), and Anhedonic Depression (depression) related to their corresponding “gold-standard” in-lab measure. Additional exploratory real-world measures of intrinsic and extrinsic motivation and passive activity were also collected. We hypothesized that these novel real-world measures of mood and anxiety symptoms would be significantly associated with their corresponding in-lab measure and would show high variability over time in the real-world.

## Methods

### Participants

Adult volunteer research participants (ages 18-75) were recruited from the greater New York City area through the Depression and Anxiety Center at the Icahn School of Medicine at Mount Sinai (ISMMS). Participants in the MA group were included if they met DSM-V criteria for MDD, post-traumatic stress disorder (PTSD), or an anxiety disorder (including GAD, Social Anxiety Disorder, and Panic Disorder) as determined by the Structural Clinical Interview for DSM-V Axis Disorders (SCID) (American Psychiatric Association, 2013). Healthy control participants free from any current or past psychiatric diagnoses as determined by the SCID or the MINI were also enrolled. Participants were excluded if they did not speak English or own a smartphone that could run the study applications. After screening, the full MASQ was completed in-lab to assess Anxious Arousal, General Distress, and Anhedonic Depression (Watson et al., 1995). All study procedures were conducted in accordance with the guidelines and regulations set by the Program for Protection of Human Subjects and Institutional Review Board at the ISMMS. Participants provided written informed consent and were compensated for their time.

### Digital Phenotyping

The smartphone application, *mindLAMP*, was utilized to capture active data and passive data on both Apple and Android personal smartphone devices over a 30-day study period. Active data included daily single-item measures of Anxious Arousal (anxiety), General Distress (distress), and Anhedonic Depression (depression) (see **Table 1**). These novel single-item scales were developed by summarizing questions that constitute the MASQ (Casillas and Clark, 2000) tripartite subscores for Anxious Arousal, General Distress, and Anhedonic Depression (see **Table 1**). Participants also completed two exploratory novel measures of intrinsic and extrinsic motivation on the same daily basis (see **Table 1**). The intrinsic motivation measure was developed by summarizing themes from the interest/enjoyment subscale of the Intrinsic Motivation Inventory (“Intrinsic Motivation Inventory (IMI) – selfdeterminationtheory.org,” n.d.) and the Work Extrinsic and Intrinsic Motivation Scale (Tremblay et al., 2009). Meanwhile the extrinsic motivation measure was developed by summarizing themes surrounding work motivation from the Work Extrinsic and Intrinsic Motivation Scale (Tremblay et al., 2009). Participants provided responses to each measure on a 0-10 point Likert scale, with 0 indicating ‘Strongly Disagree’ and 10 indicating ‘Strongly Agree.’ Passive data were also collected continuously in the background via smartphone sensors and served to monitor screentime and steps taken per day through a pedometer and Apple Health application (“iOS - Health,” n.d.). Participants were guided through download of *mindLAMP* onto their own smartphone. Each week, the research team assessed adherence and provided feedback to the participant via a standardized email template to encourage adherence (Currey and Torous, 2023).

**Table 1.**
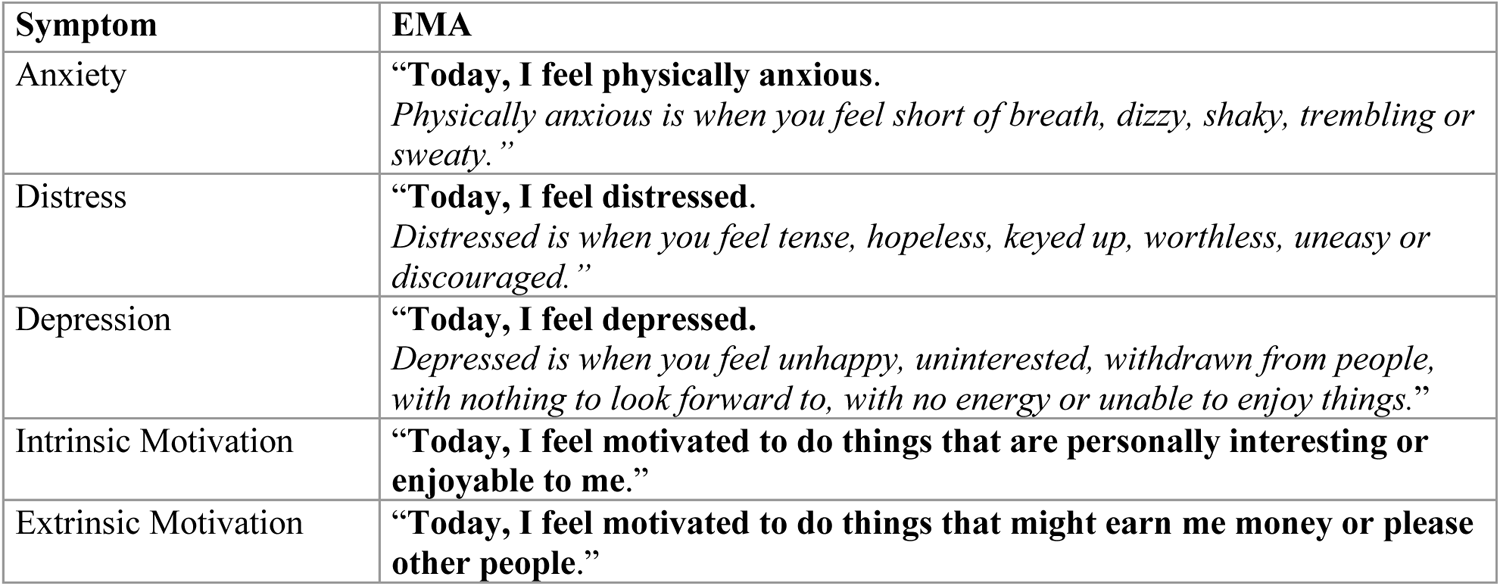
EMAs administered to participants via the *mindLAMP* application. Measures of real-world anxiety, distress, and depression were developed based on the MASQ(Watson et al., 1995). Measures of intrinsic and extrinsic motivation were developed by summarizing themes from the Intrinsic Motivation Inventory and the Work Extrinsic and Intrinsic Motivation Scale(Ryan, 1982; Ryan et al., 1983; Tremblay et al., 2009), respectively.

| Symptom | EMA |
| --- | --- |
| Anxiety | **“Today, I feel physically anxious.**<br>*Physically anxious is when you feel short of breath, dizzy, shaky, trembling or sweaty.”* |
| Distress | **“Today, I feel distressed.**<br>*Distressed is when you feel tense, hopeless, keyed up, worthless, uneasy or discouraged.”* |
| Depression | **“Today, I feel depressed.**<br>*Depressed is when you feel unhappy, uninterested, withdrawn from people, with nothing to look forward to, with no energy or unable to enjoy things.”* |
| Intrinsic Motivation | **“Today, I feel motivated to do things that are personally interesting or enjoyable to me.”** |
| Extrinsic Motivation | **“Today, I feel motivated to do things that might earn me money or please other people.”** |

### Preprocessing

Active and passive data were preprocessed in *Python.* Days were stratified to begin at 6am to account for potential duplicates from the built-in sensors. A flow chart illustrating the breakdown of participants is shown in **Figure 1**. Briefly, while N=114 participants were enrolled in the study, N=8 participant’s data were unable to be preprocessed due to technical issues resulting in no data, or a complete lack of participant engagement. Therefore N=106 participants data were successfully downloaded and preprocessed. Of these, N=101 participants (49 HC, 52 MA) met criteria for inclusion based on completing at least 3 days of active data (see **Table 2** for participant demographics). These participants were included in an adherence analysis given the study requirements for completing 30 days of surveys. Of the N=52 participants in the MA group with active data, N=47 had MASQ scores available and were included in the analysis for assessing the reliability of the real-world scales.

**Figure 1.**
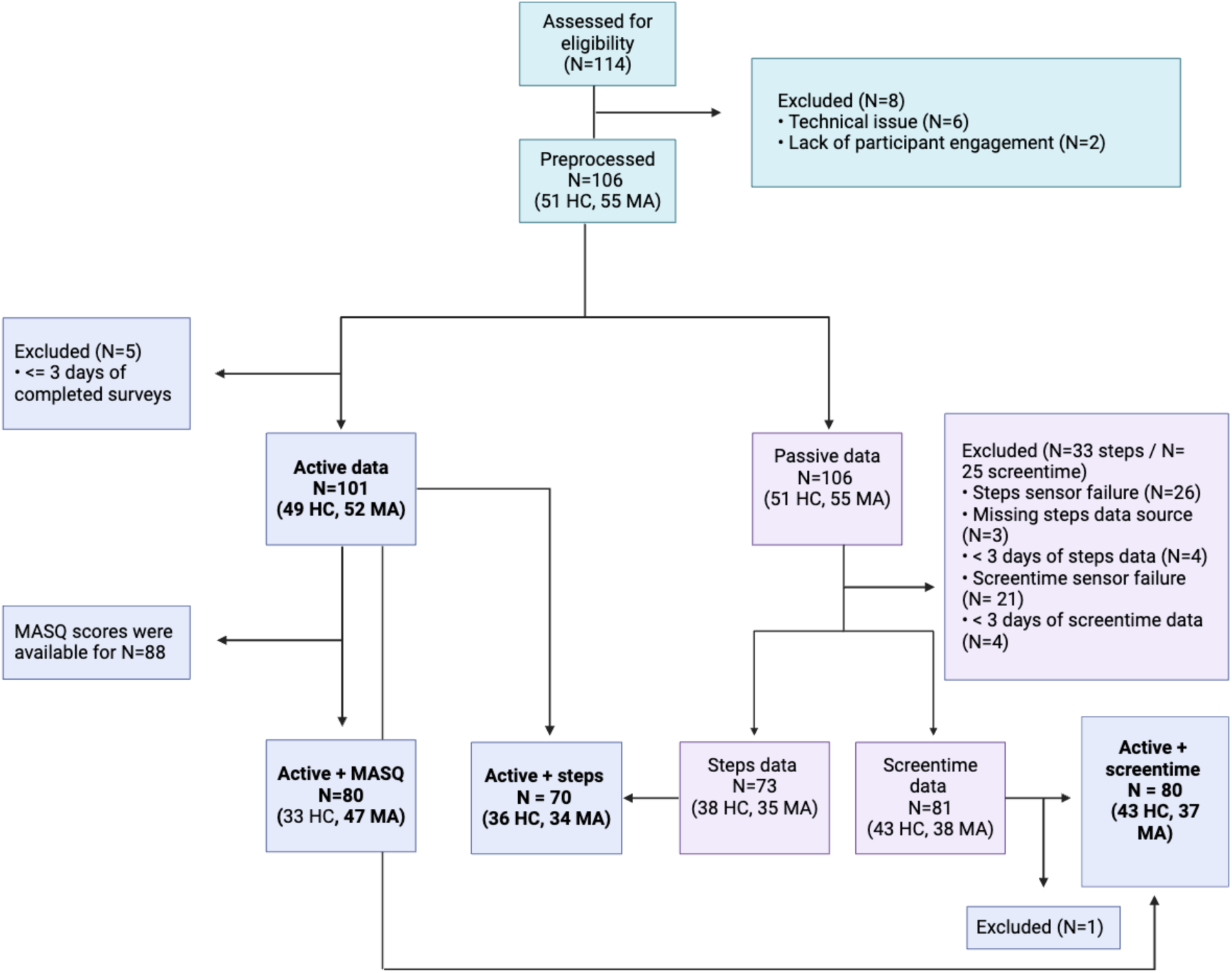
Flowchart illustrating phases of data preprocessing and participants included in each set of analyses. Bold text indicates datasets in which analyses were performed*. HC = Healthy Control group, MA = Mood/Anxiety disorder group*

**Table 2.** Demographics reported for healthy controls (HC) and participants with a mood or anxiety disorder (MA) (N= 32 MDD, 15 GAD, 5 PTSD). Welch two-sample t-tests were used to examine differences between continuous variables while Chi-squared tests examined differences between categorical variables. For race, income, and education, we report counts and percentages for the most common category.

| Participant Demographics and Clinical features |  |  |  |
| --- | --- | --- | --- |
| | HC (N= 49)<br>mean/count (SD/%) | MA (N= 52)<br>mean/count (SD/%) | Group Comparison t/<br>$\chi^2$ (p-value) |
| Age (years) | 31 (10.75) | 31.82 (9.85) | 0.383 (p=0.702) |
| Sex (female) | 30 (61.0%) | 33 (63.5%) | 6.9 x 10<sup>-4</sup> (p=0.98) |
| Race (White or Caucasian) | 22 (44.9%) | 19 (36.5%) | 0.22 (p=0.64) |
| Ethnicity (Hispanic or Latino) | 9 (18.4%) | 16 (30.8%) | 1.96 (0.162) |
| Income (\$50,001 - \$100,000) | 21 (42.9%) | 19 (36.5%) | 0.1 (0.752) |
| Education (Graduated 4-year college) | 17 (34.7%) | 12 (23.1%) | 0.862 (0.353) |
| MASQ Anxious Arousal | -- | 25.7 (9.62) | -- |
| MASQ General Distress | -- | 22.4 (9.78) | -- |
| MASQ Anhedonic Depression | -- | 39.1 (7.43) | -- |

A total of N=106 participants had passive data available upon download. However, only N=73 participants (38 HC, 35 MA) had steps sensor data and N=81 participants (43 HC, 38 MA) had screentime sensor data and met criteria for inclusion based on having at least 3 days of data. Steps sensor data was collected from two distinct sources via *mindLAMP*: 1) a pedometer within the *mindLAMP* platform which was used to calculate daily steps by taking the maximum step count on a given day and 2) Apple Health which was used to calculate steps taken per day by taking the cumulative sum of step counts on a given day. Apple Health was selected as the primary source of steps data. To account for days with missing steps data, we interpolated values from the pedometer source on days where this source had data available given the moderate correlation that exists across values between these two data sources (see **Supplementary Figure 1**). Finally, to account for a sensor-related error-margin, values recorded within 30 seconds of each other and within 10% magnitude of each other were considered duplicates and only the first entry was included (see **Supplementary Figure 2**).

Screentime sensor data were subset into epochs of morning (6am-12pm), afternoon (12pm-6pm), evening (6pm-12am), and overnight (12am-6am). The raw screentime data consisted of timestamps corresponding to each instance of change in “device state,” including “screen on,” “screen off,” “device locked,” and “device unlocked,” with each timestamp denoting the transition between these states. Preprocessing involved segmenting the data by calculating the duration between each instance of “screen on” and either “screen off” or “device locked,” accounting for instances where the device transitioned directly to “device locked.” Subsequently, the total screentime for each quadrant of the day was computed. Segments with durations of less than 30 seconds were excluded to minimize the influence of brief screen activations, often attributed to notifications rather than active use. Taking the raw data, we separately subset participants with a minimum of 5 days of active, steps, and screentime data to retain sufficient timepoints for a reliable correlation analysis between each feature. This resulted in a sample of N=48 participants (27 HC, 21 MA). Using this dataset, we extracted the time course of each of the measures to apply the Dependency Network Analysis (D_EP_NA) method described in detail in the following section (*Integrating data over time: Dependency Network Analysis (D_EP_NA)*).

### Statistical Analysis

All statistical analyses were conducted in *R.*

#### Adherence

To assess how well participants engaged with the application and determine the feasibility of digital phenotyping studies within a psychiatric patient population, we used simple, unpaired two-sample t-tests to determine if there was a significant difference in the average number of days on which participants from each group completed at least one EMA. Additionally, given the requirement to complete surveys daily for 30 days, an unpaired two-sample t-test was used to assess if there is a significant difference in survey completion by day between our two groups.

#### Assessing reliability and consistency of ecological momentary assessments

To determine the reliability of participant’s self-reported symptoms in the real-world, we assessed the relationship between in-lab MASQ scores (Anxious Arousal, General Distress, Anhedonic Depression) and their corresponding real-world score using mixed-effects models per the *lmer* package in R with: *real-world measure ∼ MASQ + day + (1 | participant)*. This reliability analysis was only conducted within the MA group given the MASQ’s specificity for detecting differences in psychiatric domains within psychiatric patient populations (Watson et al., 1995).

To determine the generalizability of our real-world scales, we constructed models with the real-world scales as the dependent variables, the corresponding MASQ score and subgroup classification (MA vs. HC) as independent variables, and participant as a random effect. Additionally, we conducted a stratified analysis to examine differences in associations across each subgroup. In this analysis, we refitted three similar regression models without the group term as a predictor for different cohorts (i.e., MA, HC, and combined MA + HC). We evaluated and visualized the differences in regression coefficients for each model using lollipop plots. Finally, to assess the internal consistency of the real-world scales over time, we extracted intraclass correlation coefficients (ICC) from the models, which were also visualized via lollipop plots. The ICC was interpreted using standard nomenclature where values below 0.5 indicate poor reliability, between 0.5 and 0.75 moderate reliability, and any value above 0.75 indicates good-to-excellent reliability (Koo and Li, 2016) (see *Supplementary Materials* for results).

All follow-up analyses were conducted in the full study sample (HC + MA). To examine variability of these three measures in the real-world, the standard deviation over time was computed for each participant in each group and entered into independent-samples t-tests, or welch tests where appropriate.

#### Exploring the effects of intrinsic and extrinsic motivation on anxiety, distress, and depression

To assess the relationship between intrinsic and extrinsic motivation and real-world symptoms in the whole sample (HC and MA), six distinct Zero-inflated Poisson (ZIP) mixed-effects models were constructed given that healthy controls consistently reported on the lower end of our real-world scales for anxiety, distress, and depression (**Supplementary Figure 3**).

ZIP models are mixture models that consist of two parts: 1) a Poisson count model which serves to estimate the incident risk ratio (IRR) and 2) a logit model for estimating an odds ratio and predicting excess zeros (“Zero-Inflated and Two-Part Mixed Effects Models,” n.d.; “Zero-Inflated Poisson Regression | R Data Analysis Examples,” n.d.). Therefore, models were specified and fitted using the GLMMadaptive package’s zi.poisson() family in R such that the count model was specified as: *real-world measure ∼ motivation + day + group + (1|participant)* while the logit model was specified as: *∼motivation* and varied in predictor variables to minimize residuals. The ZIP models specified in this analysis were selected based on lowest AIC and by assessing the normality of residuals (see *Supplementary Materials* for tables and figures).

#### Exploring the effects of physical activity and screentime on symptoms

Similarly to the motivation analysis, ZIP models were selected to separately assess the relationship between variability in real-world symptoms and physical activity (steps taken per day) and real-world symptoms and screentime within the whole sample. Count models were specified as follows for steps: *real-world measure ∼ steps_scaled + day + group + (1|participant)* and logit model *∼ steps_scaled.* For screentime the count model was specified as: *real-world measure ∼ screentime_scaled + day + group + (1|participant)* and logit model *∼ screentime_scaled.* For each of these models, the logit model varied in predictor variables to minimize residuals (see *Supplementary Materials* for tables and figures).

#### Integrating data over time: Dependency Network Analysis (D_EP_NA)

To assess how all measures interact with one another over time, we applied the D_EP_NA model to the full dataset (Jacob et al., 2018a, 2018b, 2016; Yael Jacob et al., 2019; Y. Jacob et al., 2019; Kenett et al., 2010, 2011; Madi et al., 2011). D_EP_NA is a graph theory network method for constructing a directed graph and estimating the direction of influence among nodes within the whole network, where each node is an individual digital phenotyping variable such as depression symptom severity or screentime. D_EP_NA quantifies the influence of each network node according to its partial correlation influence. This analysis, described in detail elsewhere (Jacob et al., 2016), quantifies the impact of a node over the connectivity of other pairs of nodes. D_EP_NA offers a new computational model for quantifying and comparing directed graphs based on timeseries data.

In this approach, each of the active and passive data measures represents a node in the graph. First, all the nodes (i.e. measures) time course were normalized using Z-Score. Then, the pairwise node– node connectivity matrix was calculated using Pearson correlations and normalized using a Fisher r-to-Z transformation. We then define the influence of node j on the pair of elements i and k as the difference between the correlation and the partial correlation, given by the following equation:

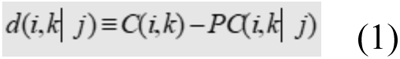

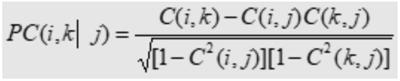

This quantity is large only when a significant fraction of the correlation between nodes i and k can be explained in terms of node j. We then calculate the partial correlation effect for each node on all other pairwise correlations in the network. The total influence of node j on node i, D(i,j) is defined as the average influence of node j on the correlations C(i,k), over all nodes k, given by:

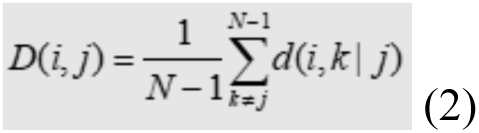

The node dependencies define a dependency matrix D, whose (i,j) element is the influence of node j on node i. Particularly, the dependency matrix is nonsymmetrical since the influence of node j on node i is not equal to the influence of node i on node j.

The ‘Influencing Degree’ of node j is defined as the sum of the influence of node j on all other nodes i, that is:

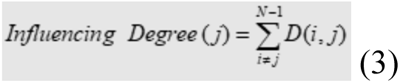

The ‘Influencing Degree’ measure indicates the hierarchy of efferent (out-degree) influence of the node on the entire network. The higher this measure, the greater its impact on all other connections in the network and the more likely it is to generate the information flow in the network. The influence of the network on node j is termed the ‘influenced degree’ and is defined as the sum of the influences (or dependencies) of all other nodes i in the network on node j, that is:

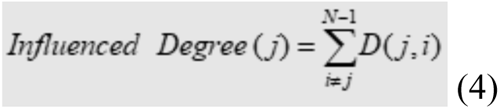

The higher the ‘Influenced Degree’ measure the more this node was dependent or influenced by all the other nodes in the network.

Next, we conducted a between-group two-sample t-test for each node’s degree of influence. All results were corrected for multiple comparisons using false discovery rate (FDR) correction with p < 0.05 threshold. To create network graph visualization, we used the pair-wise dependency connectivity matrix. A two-tailed t-statistic was computed to compare the two groups. We then connected only pair-wise nodes with dependencies that were significantly different between the two groups (p < 0.05, FDR corrected for number of nodes) creating a simple graph visualization of the differences between the groups. Graph visualization was conducted using the *NetworkX* library in Python (Hagberg et al., 2008).

## Results

### Participants

A total of N=114 participants participated in the study. Following data cleaning and preprocessing (see *Methods: Preprocessing*, **Figure 1**), a total of N=101 participants remained, including N=49 HC and N=52 in the MA group. The MA group included N=32 individuals with primary MDD and N=20 with an anxiety or stress-related disorder (see **Table 2**).

### Adherence

Over the 30-day period, HCs completed an average of 21.3 ± 6.23 total days of EMAs while the MA group completed an average of 20.9 ± 5.72 total days, with no significant difference in days of surveys completed between groups (**Figure 2A**, t_(96.9)_ = 0.403, p = 0.687). With respect to whether there was a significant difference in survey completion by day between the two groups, an average of 71.2% of HC and 69.5% of MA completed surveys each day, with no significant difference in adherence between groups (**Figure 2B**, t_(58)_ = 0.874, p=0.386), suggesting an overall good adherence and in line with prior EMA studies conducted in participants with MDD showing completion rates ranging from 65% - 85% (Jones et al., 2021).

**Figure 2.**
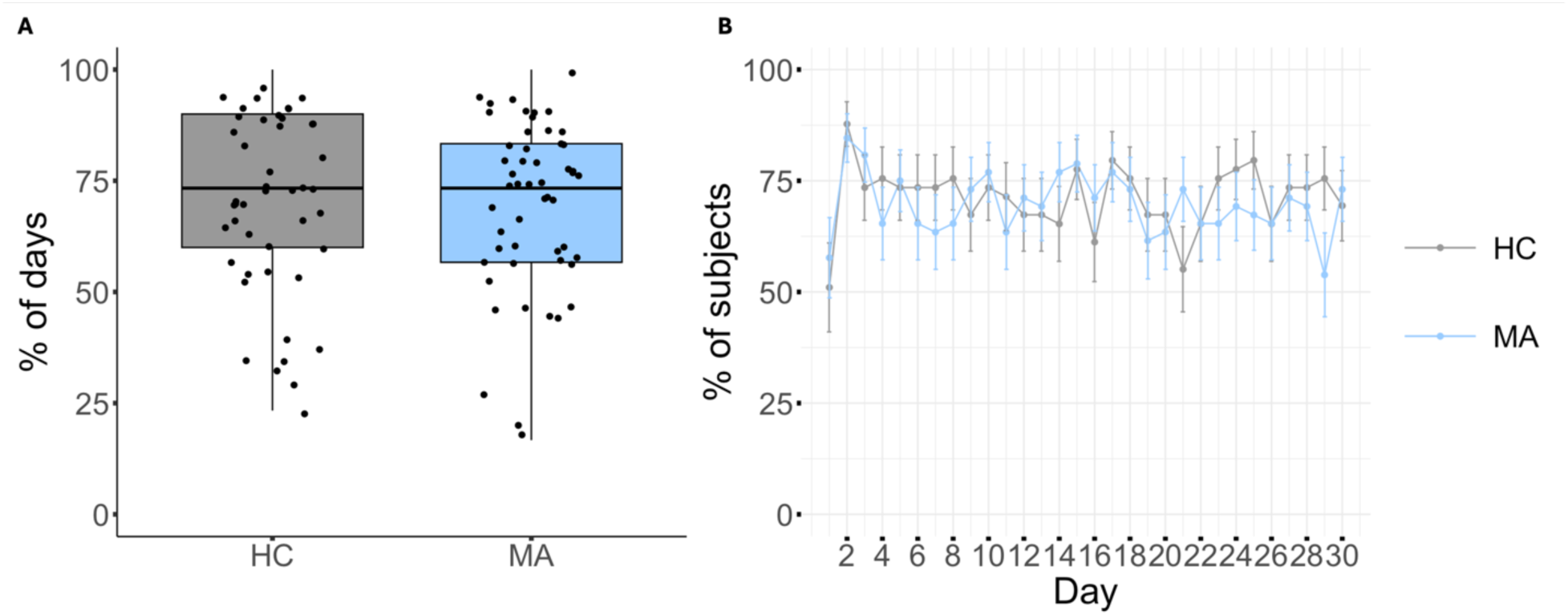
Study Adherence between groups. **A.** Box plots illustrate the percentage of days out of the 30-day requirement on which participants completed at least one survey. **B**. Line graph illustrates the percentage of subjects who completed at least one survey on a given day out of the total number enrolled to determine study adherence. *Error bars = standard deviation of a proportion, HC = Healthy Control group, MA = Mood/Anxiety disorder group*

### Relationships between real-world and in-lab measures of symptom severity

Using a linear mixed-effects model adjusted for time and individual, we found that in the MA group, daily real-world single–item measures for anxiety, distress, and depression were associated with their corresponding in-lab measures (**Figure 3B-D**, MASQ Anxious Arousal, t_(46.9)_ = 2.33, p = 0.024; General Distress, t_(46.4)_ = 4.65, p < 0.001; Anhedonic Depression, t_(46.9)_ = 2.73, p =0.009). The variability of all three real-world measures was higher in the MA group compared to the HC group (anxiety, t_(99)_ = - 6.09, p < 0.001; distress, t_(99)_ = -5.70, p < 0.001; depression, t_(87.1)_ = -4.99, p < 0.001) (see Supplementary Figure 4).

**Figure 3.**
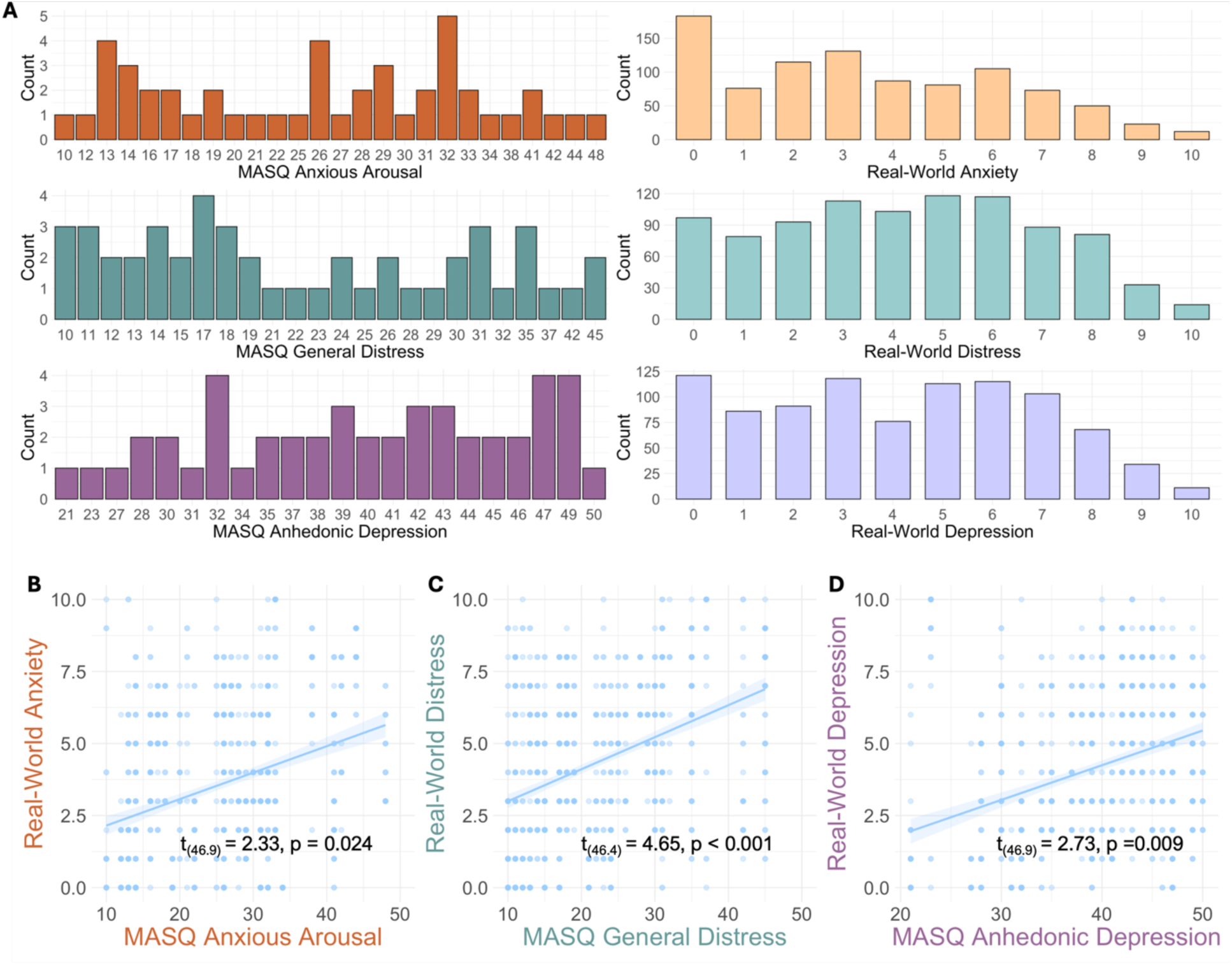
Results from assessing the reliability of real-world measures for anxiety, distress, and depression within the MA group. **A.** Distribution of the MA group’s MASQ scores beside their corresponding anxiety, distress, and depression EMA score distributions. **B-D.** Linear mixed-effects regression models showcasing significant relationships between the in-lab MASQ for Anxious Arousal, General Distress, and Anhedonic Depression against real-world anxiety, distress, and depression scores (t_(46.9)_ = 2.33, p = 0.024; t_(46.4)_ = 4.65, p < 0.001; t_(46.9)_ = 2.73, p =0.009). Individual data points represent survey responses per participant. *MA = Mood/Anxiety disorder group, MASQ= Mood and Anxiety Symptom Questionnaire*

In the full cohort (i.e., MA + HC), there was a significant association between the MASQ Anxious Arousal and real-world anxiety after adjusting for day and group (**Figure 4**, t_(1565)_= 4.61, p < 0.001). Associations were also seen in the stratified analyses for MA (t_(909)_ = 2.28, p = 0.023) and HC groups (t_(656)_ = 6.46, p < 0.001). Similarly, in the full cohort, a significant association was observed between the MASQ General Distress and real-world distress after adjusting for day and group (**Figure 4**, t_(1567)_ = 6.50, p < 0.001). Associations were also seen in the stratified analyses for MA (t_(909)_ = 4.55, p < 0.001) and HC groups (t_(657)_ = 6.94, p < 0.001). Finally, in the full cohort, a significant association was observed between the MASQ Anhedonic Depression and real-world depression after adjusting for day and group (**Figure 4**, t_(1553)_ = 3.21, p < 0.001). Associations were also seen in the stratified analyses for MA (t_(901)_ = 2.67, p = 0.008) and HC (t_(651)_ = 2.03, p = 0.043) groups. See *Supplementary Materials* for additional results for the association between the MASQ total score and real-world anxiety/distress/depression total score.

**Figure 4.**
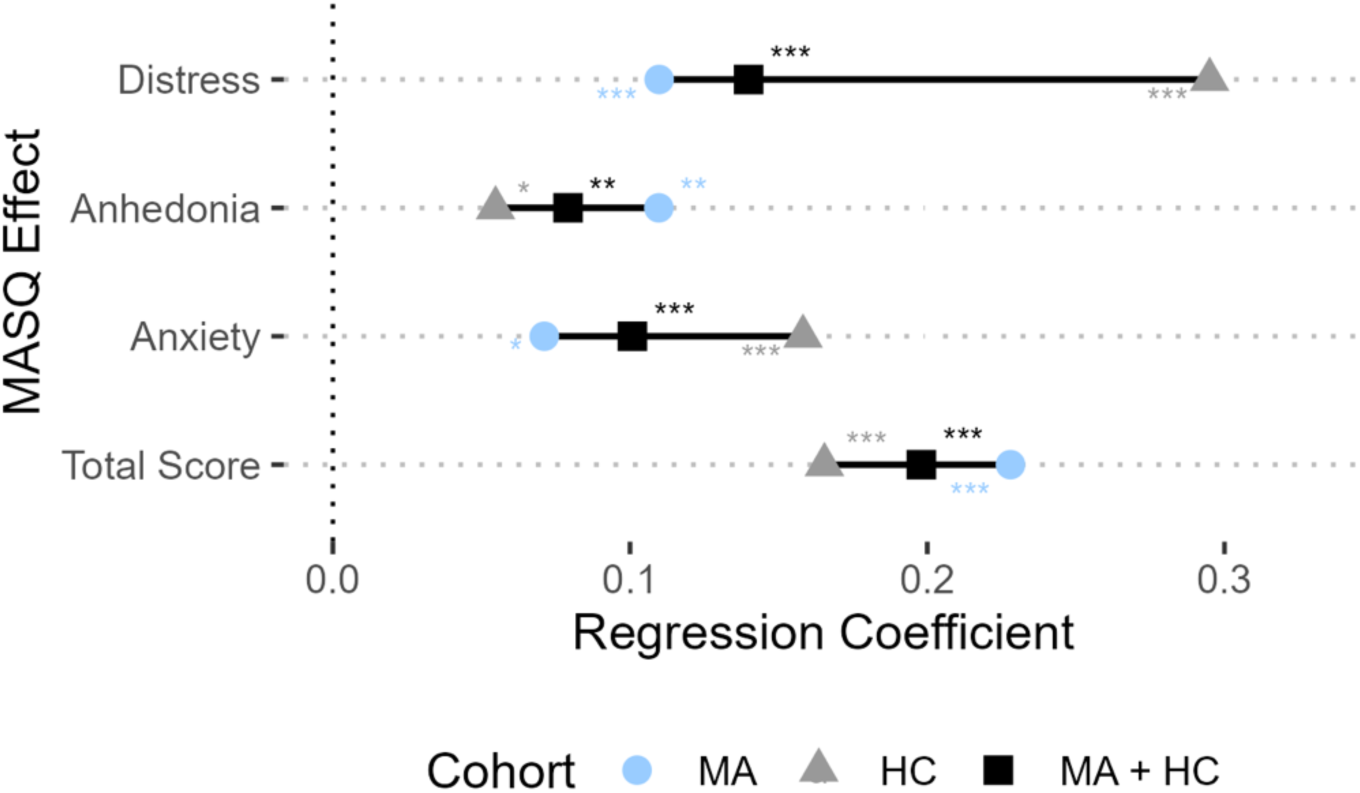
Results from assessing the generalizability of real-world measures for anxiety, distress, and depression. Lollipop plots depict the regression coefficient for the MASQ subscale scores in relation to their corresponding real-world scales between the full cohort (HC + MA) and each subgroup. In the full cohort, significant associations were found between the MASQ Anxious Arousal/General Distress/Anhedonic Depression and their corresponding real-world scale demonstrating the generalizability of these scales (t’s >2.28, p’s < 0.05). *HC = Healthy Control, MA = Mood/Anxiety disorder group*

**Figure 5.**
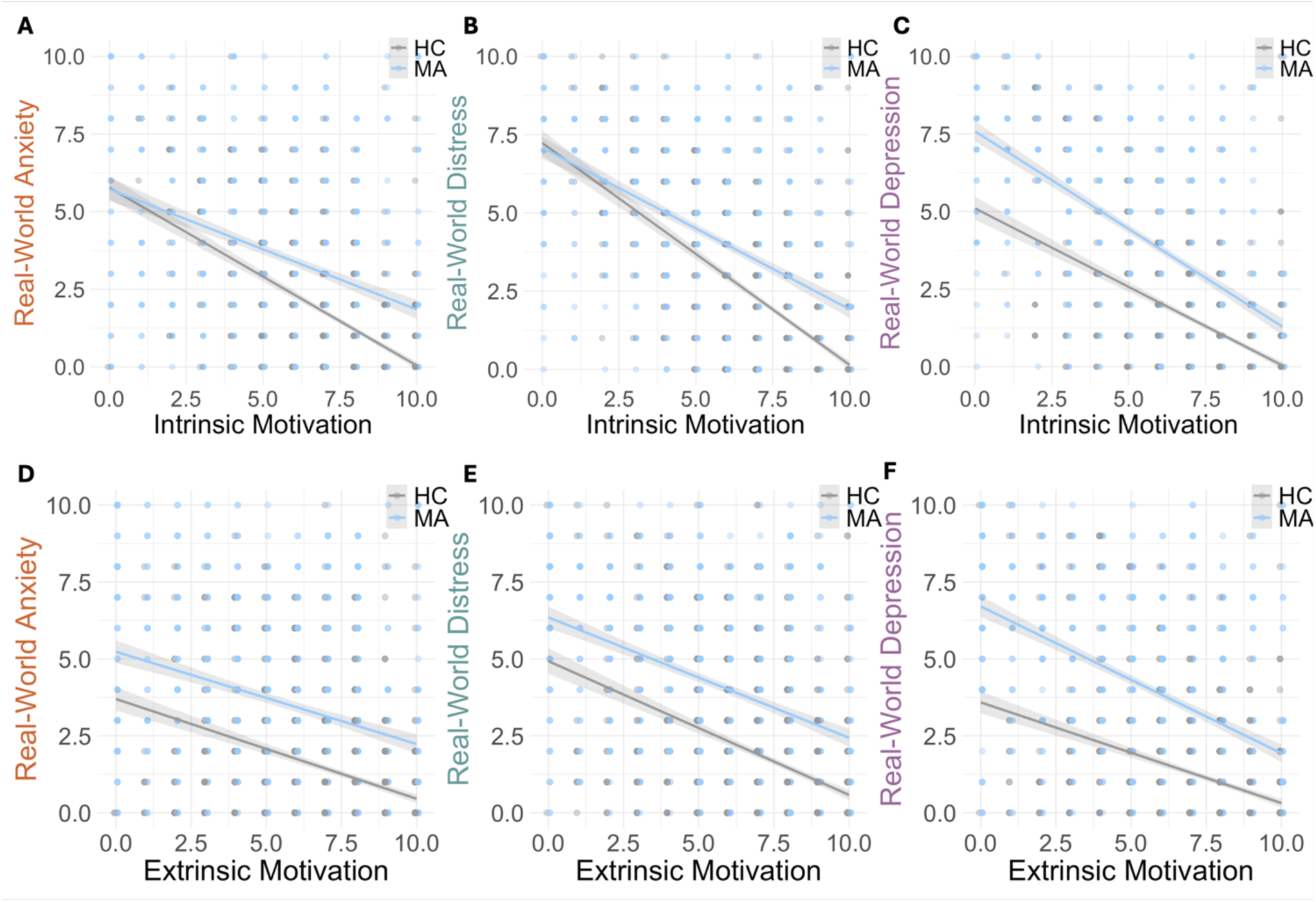
ZIP regression models illustrating the relationship between intrinsic **(A-C)** and extrinsic **(D-F)** motivation and real-world anxiety, distress, and depression. For each anxiety/distress/depression model there were main effects of intrinsic motivation (IRRs > 0.82, p’s < 0.001) and extrinsic motivation (IRRs > 0.88, p’s ≤0.001). There were also significant interactions between group and intrinsic motivation on anxiety/distress/depression (IRRs > 1.08, p’s < 0.001) and group and extrinsic motivation on depression (IRR = 1.06, p = 0.019). *HC = Healthy Control group, MA = Mood/Anxiety disorder group*

### Exploring the effects of intrinsic and extrinsic motivation on anxiety, distress and depression

We assessed the relationship between real-world measures of intrinsic (**Figure 4A-C**) and extrinsic (**Figure 4D-F**) motivation, and real-world anxiety, distress, and depression on an exploratory basis. See *Supplementary Materials* for tables, model residuals, and Q-Q plots.

### Intrinsic Motivation

For anxiety, there was a main effect of intrinsic motivation on anxiety in the count regression model (IRR = 0.87, CI = (0.83-0.90), p < 0.001), whereby lower intrinsic motivation was associated with higher anxiety severity. There was also a significant interaction between intrinsic motivation and group (IRR = 1.12, CI= (1.07-1.17), p < 0.001), whereby as intrinsic motivation increased, groups differed more in anxiety severity. There was no significant effect of group (IRR = 1.23, CI = (0.83-1.83), p = 0.303), or day (IRR = 1.00, CI = (1.00-1.00), p = 0.447). The logit model similarly revealed a main effect of intrinsic motivation on anxiety (IRR = 1.62, CI = (1.29 -2.04), p < 0.001), whereby the odds of observing zero symptoms of anxiety increased with higher intrinsic motivation. There was no significant effect of group in the logit model (IRR = 0.20, CI= (0.03 - 1.58), p = 0.127).

For distress, there was a main effect of intrinsic motivation on distress in the count regression model (IRR = 0.86, CI = (0.83-0.89), p <0.001), whereby lower intrinsic motivation was associated with a higher distress severity. There was also a significant interaction between intrinsic motivation and group on distress (IRR = 1.08, CI = (1.04 - 1.12), p < 0.001), whereby as intrinsic motivation increased, groups differ more in distress severity. Again, there was no significant effect of group (IRR = 1.21, CI = (0.87-1.69), p= 0.260), or day (IRR = 1.00, CI = (1.00-1.00), p= 0.941). The logit model similarly revealed main effects of intrinsic motivation on presence of distress (IRR = 2.67, CI = (1.85 - 3.85), p <0.001) and day on distress (IRR = 1.06, CI = (1.02-1.10), p = 0.003), whereby the odds of observing zero symptoms of distress increased with higher intrinsic motivation and by day. This effect varied by group such that participants in the MA group had lower odds of zero distress symptoms compared to HCs (IRR = 0.110, CI = (0.02-0.69), p = 0.019).

For depression, there was also a main effect of intrinsic motivation on depression in the count regression model (IRR = 0.82, CI = (0.79 - 0.86), p < 0.001), whereby lower intrinsic motivation was associated with higher depression severity. There was a significant interaction between intrinsic motivation and group on depression (IRR = 1.11, CI = (1.06 - 1.17), p < 0.001), whereby as intrinsic motivation increased, groups differed more in depression severity. There was no significant effect of group (IRR = 1.10, CI = (0.76 -1.58), p =0.629), or day (IRR = 1.00 CI = (1.00-1.00), p = 0.886). The logit model similarly revealed a main effect of intrinsic motivation on presence of depression (IRR = 2.29, CI = (1.60 – 3.29), p <0.001), as well as day (IRR = 1.05, CI = (1.01-1.10), p = 0.013), whereby the odds of observing zero symptoms of depression increased with higher intrinsic motivation and by day. This effect varied by group such that participants in the MA group had lower odds of zero depression symptoms compared to HCs (IRR = 0.01 CI = (0.00-0.16), p < 0.001).

#### Extrinsic Motivation

For anxiety, there was a main effect of extrinsic motivation on anxiety in the count regression model (IRR =0.93, CI = (0.90-0.99), p = 0.001), whereby lower extrinsic motivation was associated with a higher anxiety severity. There was a significant interaction between extrinsic motivation and group on anxiety (IRR = 1.05, CI= (1.01 – 1.10), p = 0.024), whereby as extrinsic motivation increased, groups differed more in anxiety severity. There was also a main effect of group on anxiety (IRR = 2.07, CI= (1.39-3.08), p <0.001), whereby participants in the MA group experience greater anxiety compared to HCs, with no effect of day (IRR = 1.00, CI = (1.00-1.00), p= 0.676). The logit model similarly revealed a main effect of extrinsic motivation on presence of anxiety (IRR = 1.62, CI = (1.27 – 2.07), p < 0.001), whereby the odds of observing zero symptoms of anxiety increases with higher extrinsic motivation. There was no significant effect of group (IRR = 0.15, CI = (0.02-1.23), p = 0.077).

For distress, there was a main effect of extrinsic motivation on distress (IRR = 0.93, CI = (0.98 – 0.97), p <0.001), whereby lower extrinsic motivation was associated with a higher distress severity. There was a main effect of group on distress (IRR = 2.02, CI = (1.42 – 2.87), p < 0.001), whereby participants in the MA group experience greater distress compared to HCs, with no effect of day (IRR = 1.00, CI = (1.00-1.00), p= 0.431), and no significant interaction between extrinsic motivation and group on distress (IRR = 1.02, CI= (0.98 – 1.06), p = 0.297). The logit model similarly revealed a main effect of extrinsic motivation on presence of distress (IRR = 2.22, CI = (1.54 – 3.19), p < 0.001), whereby the odds of observing zero symptoms of distress increases with higher extrinsic motivation by day (IRR = 1.05, CI = (1.01-1.09), p = 0.009). This effect varies by group such that participants in the MA group have a decrease in the odds of observing zero distress symptoms compared to HCs (IRR = 0.09, CI = (0.01-0.57), p = 0.010).

For depression, there was also a main effect of extrinsic motivation (IRR = 0.88, CI = (0.85 – 0.92), p < 0.001) and day on depression (IRR=1.00, CI=(0.99-1.00), p=0.059), whereby greater extrinsic motivation was associated with lower depression severity and a decrease in depression symptoms over time. There was also a main effect of group on depression (IRR = 1.70, CI = (1.16 – 2.48), p = 0.007), whereby participants in the MA group experience greater depression symptoms compared to HCs. There was also an interaction between group and extrinsic motivation on depression (IRR = 1.06, CI = (1.01 – 1.10), p = 0.019), whereby as extrinsic motivation increases, groups differ more in depression severity. The logit model similarly revealed a main effect of extrinsic motivation on presence of depression (IRR = 1.33, CI = (1.06 – 1.65), p= 0.012), whereby the odds of observing zero symptoms of depression increases with higher extrinsic motivation. Again, this effect varies by group such that participants in the MA group have a decrease in the odds of observing zero depression symptoms compared to HCs (IRR = 0.01, CI = (0.00 – 0.06), p < 0.001).

### Exploring the effects of activity on anxiety, distress and depression

#### Physical Activity: Steps

For anxiety (**Figure 6A**), there was no main effect of steps or day on anxiety severity in the count regression model (steps, IRR = 0.98, CI = (0.94-1.03), p=0.46; day, IRR = 1.00, CI = (1.00-1.00), p=0.939). There was a main effect of group on anxiety (IRR = 3.28, CI = (2.29-4.69), p < 0.001), whereby participants in the MA group reported greater anxiety severity compared to HCs. Similarly, the logit model showed a main effect of group (IRR = 0.06, CI = (0.00-1.03), p = 0.052) and there was no effect of steps (IRR = 1.09, CI = (0.69-1.73), p= 0.716) or day (IRR = 0.98, CI = (0.93-1.03), p = 0.478) on anxiety.

**Figure 6.**
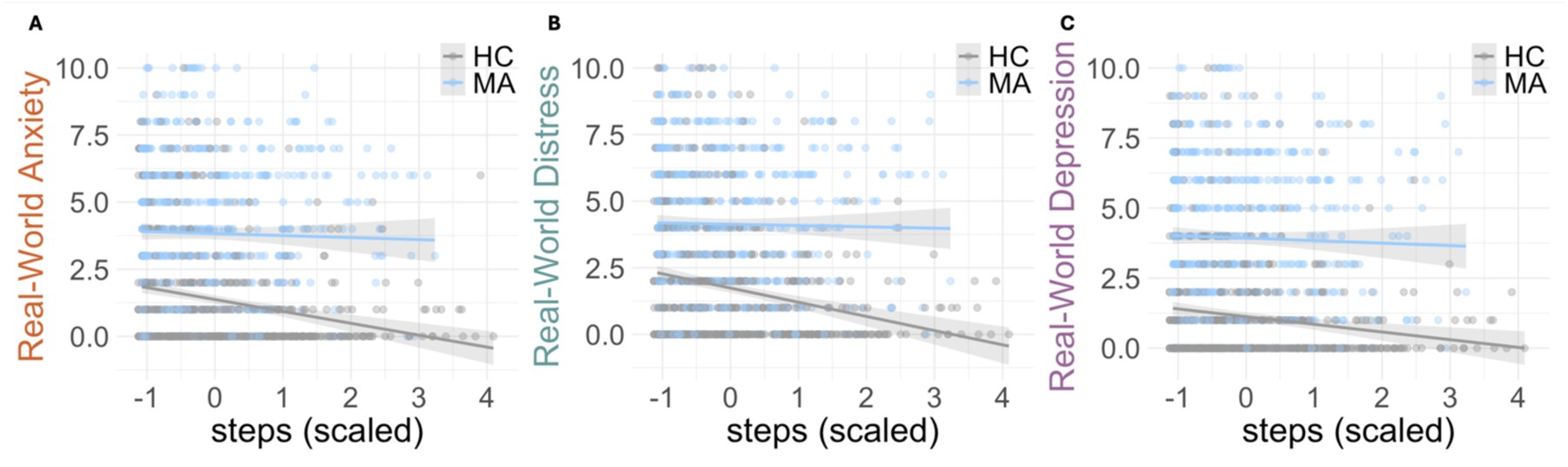
Relationship between steps and real-world anxiety, distress, and depression. A main effect of steps on real-world distress and depression (but not anxiety) was observed (IRRs > 0.93, p’s ≤ 0.05). Across all three measures, participants in the MA group experience greater symptom severity (IRRs > 2, p’s < 0.001) in comparison to HC. *HC = Healthy Control group, MA = Mood/Anxiety disorder group*

For distress (**Figure 6B**), there was a main effect of steps in the count regression model (IRR = 0.96, CI = (0.91 - 1.00), p = 0.059), whereby higher physical activity was associated with lower severity of distress. There was also a main effect of group (IRR = 2.18, CI = (1.64 - 2.92), p < 0.001) on distress, whereby participants in the MA group reported greater distress severity compared to HCs, with no effect of day (IRR = 1.00, CI = (0.99 - 1.00), p = 0.406). The logit model revealed a main effect of group (IRR = 0.02, CI = (0.00-0.13), p <0.001), such that participants in the MA group had lower odds of zero symptoms of distress. There was a main effect of day (IRR = 1.05, CI = (1.01-1.09), p=0.022) whereby the odds of observing zero symptoms of distress increased over time, but there was no effect of steps (IRR = 1.14, CI = (0.73 - 1.80), p = 0.556).

For depression (**Figure 6C**), there was a main effect of steps on depression in the count regression model (IRR = 0.93, CI = (0.89 - 0.98), p = 0.003), whereby higher physical activity was associated with lower severity of depression. There was also a main effect of group (IRR = 2.13, CI = (1.54 - 2.95), p < 0.001) and day (IRR = 0.99, CI = (0.99-1.00), p=0.025), whereby participants in the MA group reported greater depression severity compared to HCs and lower depression severity was observed over time. The logit model revealed a main effect of group (IRR = 4.6 x 10^-4^, CI = (0.00-0.01), p < 0.001) on depression, whereby participants in the MA group had lower odds of observing zero symptom severity of depression.

#### Digital Activity: Screentime

For anxiety (**Figure 7A**), there was no main effect of screentime on anxiety (IRR = 1.02, CI = (0.96 – 1.07), p = 0.573) in the count regression model. There was a main effect of day (IRR = 1.01, CI = (1.00 – 1.01), p = 0.032) and group (IRR = 2.67, CI = (1.90 – 3.78), p < 0.001), whereby participants in the MA group reported greater anxiety severity compared to HCs and lower anxiety severity observed over time. The logit model revealed no significant effects of screentime (IRR = 1.50, CI = (0.90 – 2.52), p = 0.120), day (IRR = 1.04, CI = (0.99 – 1.09), p = 0.087), or group (IRR = 0.05, CI = (0.00 – 1.65), p = 0.093) on anxiety.

**Figure 7.**
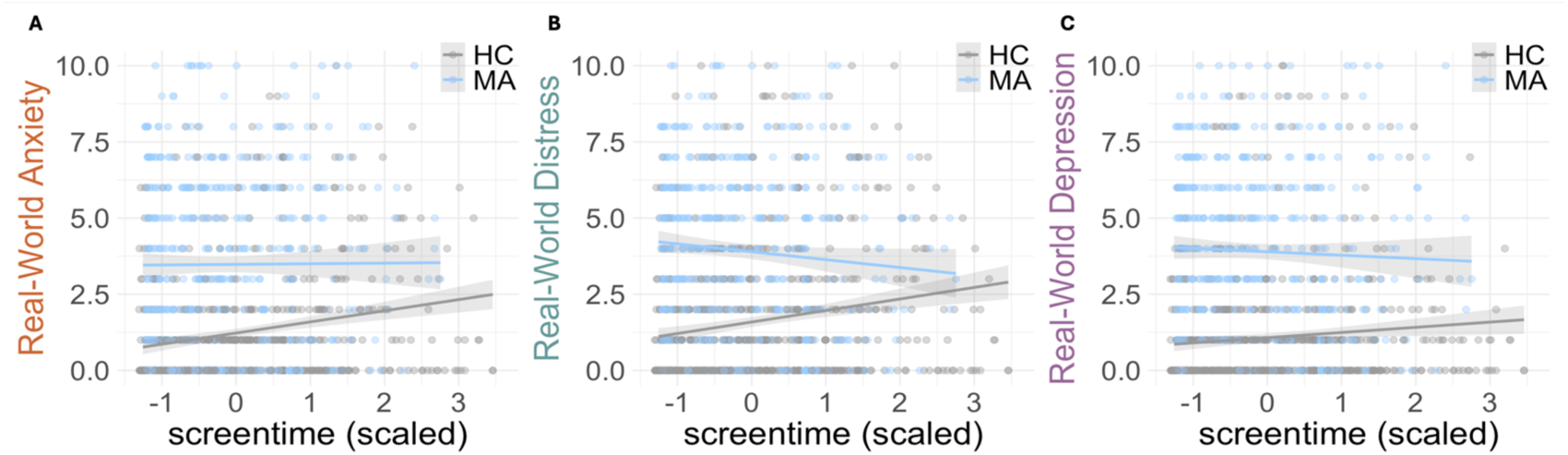
Relationship between screentime and real-world anxiety, distress, and depression. There was no effect of screentime on real-world anxiety, distress and depression in the count model (IRRs > 1.00, p’s > 0.05). Meanwhile, the logit model revealed a main effect of screentime only on distress (IRR =1.85, CI = (1.18 – 2.90), p = 0.007), whereby the odds of observing zero symptoms of distress increase with higher screentime. Across all three measures, participants in the MA group experience greater symptom severity (IRRs > 2, p’s < 0.001) in comparison to HCs. *HC = Healthy Control group, MA = Mood/Anxiety disorder group*

For distress (**Figure 7B**), there was no main effect of screentime (IRR = 1.00, CI = (0.95 – 1.05), p = 0.987) or day (IRR = 1.00, CI = (1.00 – 1.01), p = 0.311) on distress in the count regression model. There was a main effect of group on distress (IRR = 2.16, CI = (1.63 – 2.86), p < 0.001), whereby participants in the MA group reported greater distress severity compared to HCs. The logit model revealed a main effect of screentime on distress (IRR =1.85, CI = (1.18 – 2.90), p = 0.007), whereby the odds of observing zero symptoms of distress increased with higher screentime. There was also a main effect of day (IRR=1.08, CI = (1.04 – 1.13), p < 0.001), and group (IRR= 0.02, CI = (0.00, 0.25), p=0.003) on distress such that participants in the MA group had lower odds of observing zero symptoms of distress and the odds of observing zero symptoms of distress increased over time.

For depression (**Figure 7C**), there was no main effect of screentime (IRR = 1.01, CI = (0.96 – 1.07), p = 0.625) or day (IRR = 1.00, CI = (0.99 – 1.01), p = 0.851) on depression in the count regression model. There was a main effect of group on depression (IRR=2.81, CI= (2.01 – 3.92), p < 0.001), whereby participants in the MA group reported greater depression severity compared to HCs. Similarly, the logit model revealed a main effect of group (IRR = 0.00410, CI = (0.00 - 0.14), p=0.002) and day (IRR=1.11, CI = (1.03, 1.19), p = 0.008), such that participants in the MA group showed lower odds of observing zero symptoms of depression and the odds of observing zero symptoms of depression increased over time. There was no effect of screentime (IRR = 0.60, CI = (0.25 – 1.43), p = 0.249), on depression.

### Assessing interactions over time: D_EP_NA

The D_EP_NA method provided an estimation of the influence of each individual symptom measure on the entire network of symptom and activity measures over time (**Figure 8A-C**). In terms of overall influence, intrinsic motivation and extrinsic motivation had the highest *Influencing Degree* on the rest of the symptom network in the MA group (**Table 3**). Indeed, the MA group exhibited significantly higher influence of extrinsic motivation and intrinsic motivation, as compared to HC (t_(46)_ = 2.62, p < 0.02, q FDR < 0.05, *Cohen’s d* = 0.76 and t_(46)_ = 2.69, p < 0.01, q FDR < 0.05, *Cohen’s d* = 0.78 respectively) (**Table 3**, **Figure 7B**). Depression and extrinsic motivation were more influenced in the MA group compared to HC (t_(46)_ = 2.59, p = 0.01, q FDR = 0.07, *Cohen’s d = 0.75* and t_(46)_ = 2.46, p = 0.02, q FDR = 0.07, *Cohen’s d* = 0.71 (**Table 3**, **Figure 7C**), although this did not survive FDR-correction. In terms of specific, directed influence, intrinsic motivation significantly influenced steps (t_(46)_ = 3.24, p < 0.003, q FDR < 0.05, Cohen’s d = 0.94), to a greater extent among the MA group compared to HC (**Figure 7A**).

**Figure 8.**
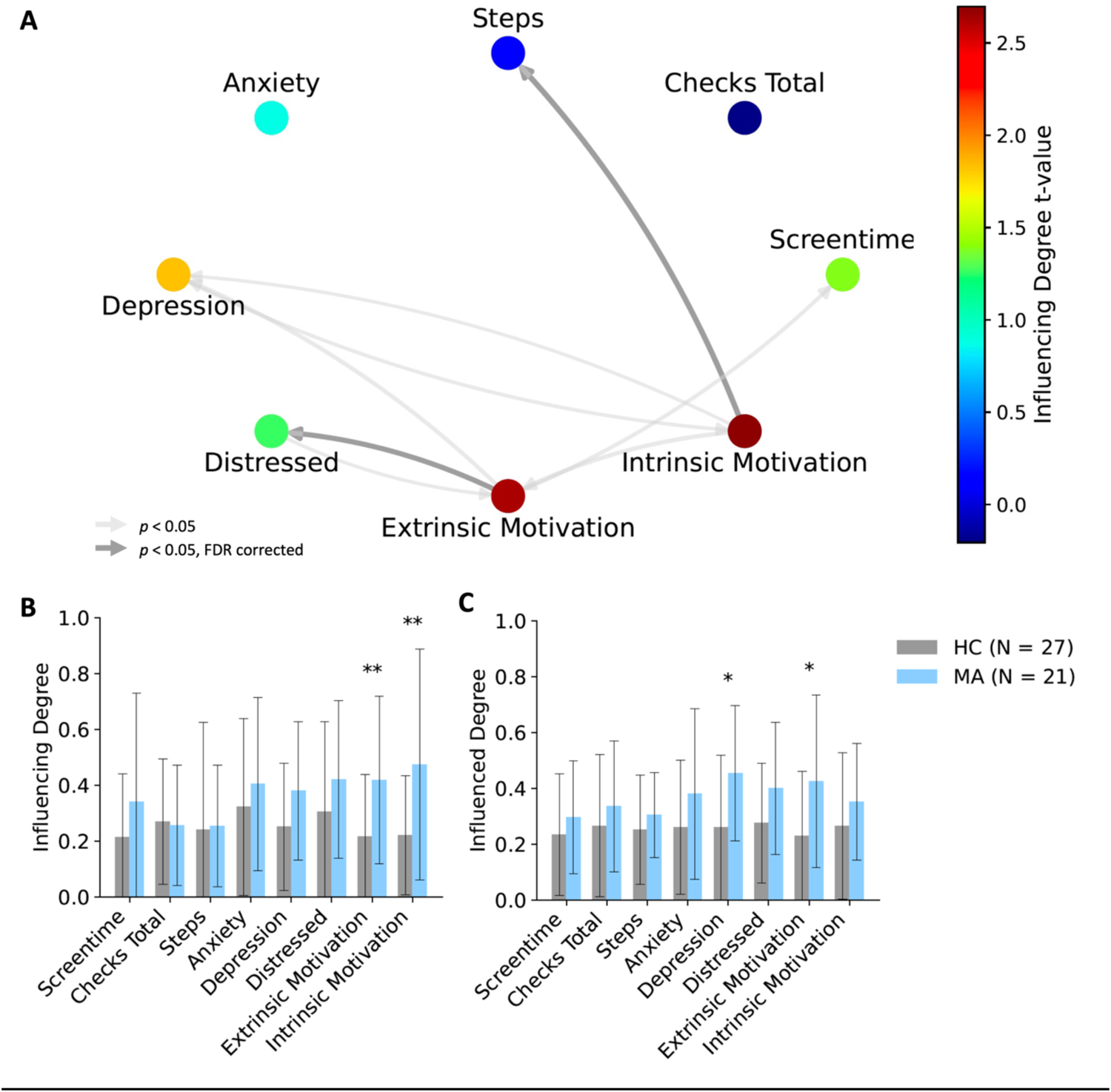
Dependency network analysis (D_EP_NA) results. **(A)** A network illustration and graph visualization of the ‘influencing degree’ of symptoms in the MA group against healthy controls. Each region is color-coded according to the t statistic value from the t-test between the ‘Influencing Degree’ of the two groups. All pair-wise ROIs with connections, significant at the p<0.05 level, are plotted as edges. **(B)** The nodes’ averaged ‘Influencing Degree’ and **(C)** ‘Influenced Degree’. The total influence of both extrinsic and intrinsic motivation was significantly higher among the MA group compared to healthy controls. *p<0.05, ** p<0.05 FDR corrected. *HC = Healthy Control group, MA = Mood/Anxiety disorder group*

**Table 3.** Influencing and influenced degree of symptoms and activity measures as estimated by the Dependency Network Analysis (D_EP_NA).

|  | *Measure* | *HC*<br>*mean ± std* | *MA*<br>*mean ± std* | *t-*<br>*value* | *p-*<br>*value* | *FDR*<br>*adjusted*<br>*p-value* | *Cohen's*<br>*d* |
| --- | --- | --- | --- | --- | --- | --- | --- |
| ***Influencing***<br>***Degree*** | Screen time | 0.21 ± 0.23 | 0.34 ± 0.39 | 1.39 | 0.17 | 0.34 | 0.40 |
|  | Checks total | 0.27 ± 0.22 | 0.26 ± 0.22 | -0.21 | 0.84 | 0.90 | -0.06 |
|  | Steps | 0.24 ± 0.38 | 0.25 ± 0.22 | 0.13 | 0.90 | 0.90 | 0.04 |
|  | Anxiety | 0.32 ± 0.32 | 0.40 ± 0.31 | 0.88 | 0.38 | 0.51 | 0.26 |
|  | Depression | 0.25 ± 0.23 | 0.38 ± 0.25 | 1.83 | 0.07 | 0.20 | 0.53 |
|  | Distress | 0.31 ± 0.32 | 0.42 ± 0.28 | 1.26 | 0.21 | 0.34 | 0.37 |
|  | Extrinsic motivation | 0.22 ± 0.22 | 0.42 ± 0.30 | 2.62 | **0.01\*** | **0.05\*** | 0.76 |
|  | Intrinsic motivation | 0.22 ± 0.21 | 0.47 ± 0.41 | 2.70 | **0.01\*** | **0.05\*** | 0.78 |
| ***Influenced***<br>***Degree*** | Screen time | 0.23 ± 0.22 | 0.30 ± 0.20 | 1.00 | 0.32 | 0.35 | 0.29 |
|  | Checks total | 0.27 ± 0.25 | 0.34 ± 0.23 | 0.95 | 0.35 | 0.35 | 0.28 |
|  | Steps | 0.25 ± 0.20 | 0.30 ± 0.15 | 0.99 | 0.33 | 0.35 | 0.29 |
|  | Anxiety | 0.26 ± 0.24 | 0.38 ± 0.31 | 1.48 | 0.15 | 0.29 | 0.43 |
|  | Depression | 0.26 ± 0.26 | 0.45 ± 0.24 | 2.59 | **0.01\*** | 0.07 | 0.75 |
|  | Distress | 0.28 ± 0.21 | 0.40 ± 0.24 | 1.87 | 0.07 | 0.18 | 0.54 |
|  | Extrinsic motivation | 0.23 ± 0.23 | 0.43 ± 0.31 | 2.46 | **0.02\*** | 0.07 | 0.71 |
|  | Intrinsic motivation | 0.27 ± 0.26 | 0.35 ± 0.21 | 1.22 | 0.23 | 0.35 | 0.36 |

On the other hand, extrinsic motivation influenced distress to a greater extent among the MA group compared to HC (t_(46)_ = 3.04, p < 0.004, q FDR < 0.05, Cohen’s d = 0.88) (**Figure 7A**). We did not find any significant FDR corrected results among HC compared to MA.

## Discussion

This study presents a new set of single-item self-reported real-world measures of anxiety, distress and depression that were significantly related to established in-lab measures of these symptom domains in individuals with mood and anxiety disorders. Novel, exploratory digital phenotyping measures of extrinsic and intrinsic motivation were also significantly related to mood and anxiety symptoms in the real-world. Both types of motivation significantly influenced all other symptom and activity measures to a greater extent in the mood and anxiety group compared to the HC group. Finally, physical activity (steps) was significantly associated with the severity of depression and distress but not anxiety; whereas digital activity (screentime) was only associated with the absence of distress. These results demonstrate how digital phenotyping measures can reliably measure symptom severity and symptom interactions in the real-world in individuals with mood and anxiety disorders.

Overall, the current results demonstrate the utility and feasibility of digital phenotyping for accurately monitoring symptoms in participants with psychiatric conditions that have been associated with high burden and drop-out rates (Torous et al., 2020, 2018). Indeed, in this work we found similarly good adherence levels between groups, suggesting that the use of few >=5 single-item daily surveys over 30-days, alongside weekly check-ins provide good adherence within a feasible, low-burden framework. Indeed, previous work demonstrates that providing some form of digital feedback can enhance adherence (Joseph et al., 2021; Mouchabac et al., 2021). Maintaining good adherence over time is critical given the significant variability in symptom severity in individuals with mood and anxiety disorders demonstrated.

Previous work assessing the relationship between extrinsic and intrinsic motivation and real-world anxiety, distress, and depression has been limited, in part due to a lack of consensus on the precise definition of intrinsic motivation, and how it can be distinguished from extrinsic motivation (Lee et al., 2012; Morris et al., 2022). Reduced *extrinsic* motivation and sensitivity to extrinsic rewards has been consistently measured in mood disorders such as MDD (Pizzagalli et al., 2009; Treadway et al., 2012; Vrieze et al., 2013; Yang et al., 2014), but the impact of *intrinsic* motivation is unclear. Here, both extrinsic and intrinsic motivation showed significant relationships with real-world anxiety, distress, and depression such that lower motivation was associated with higher symptom severity, suggesting a lack of divergence, at least in these measures. There were significant interactions between group and intrinsic motivation for all three symptom measures, whereby groups differed more in symptom severity when intrinsic motivation was high. Interestingly, intrinsic motivation seemed to have a greater impact on depression severity than extrinsic motivation (18% decrease versus 11.7% decrease). This is in line with prior work, suggesting that when individuals can effectively engage intrinsic motivational processes such as working for personal growth or for personal satisfaction, they may be more protected against depression (Ling et al., 2016). Notably, group differences were only observed in the extrinsic motivation models whereby participants with a mood or anxiety disorder showed greater anxiety, distress, and depression symptom severity in comparison to healthy controls, when extrinsic motivation was held constant. Together this suggests that the impact of intrinsic motivation is more variable based on group membership but overall, may have more protective effects on depression than extrinsic motivation. In terms of anxiety symptoms, there was a significant interaction with group, whereby as intrinsic motivation increased, groups differed more in anxiety severity. Previous work supports how the development of high intrinsic motivation in individuals with anxiety disorders might derive from maladaptive uncertainty learning that problematically drives elevated avoidance behaviors (Charpentier et al., 2017; Winch et al., 2015). Conversely in depressive phenotypes, there is evidence suggestive of a general difficulty with engaging intrinsic motivational processes (Furman et al., 2011; Mori et al., 2018; Winch et al., 2015). While this study cannot explicitly demonstrate which kinds of intrinsic or extrinsic motivators participants drew from when completing assessments, the provided examples aligned with a mix of previous reports of intrinsic and extrinsic rewards in order to capture the entire scope of the phenotype (Chew et al., 2021; Lee and Reeve, 2017; Morris et al., 2022; Reeve, 1989). It is also not clear from this study whether intrinsic versus extrinsic reward sensitivity versus motivational tone were important, as both outcome sensitivity and internal drive or vigor could differentially contribute to the self-assessment of intrinsic or extrinsic motivation. Future work should more precisely characterize the array of intrinsic and extrinsic factors that could contribute to motivation in the real-world. Nonetheless, these findings implicate the importance of assessing for the kinds of motivators individuals experience and how they may relate to affective processes.

In terms of digital activity (screentime), we did not observe an effect of digital activity on anxiety, distress, or depression in the count models. However, there was a main effect of screentime on distress in the logit model suggesting that increased screentime was associated with higher odds of not experiencing distress. While concerns regarding the psychological and cognitive impacts of screentime persist (Rast et al., 2021; Ward et al., 2017), this latter finding may be reflective of the positive impact of smartphone usage (Firth et al., 2024; Przybylski et al., 2020). Recent work emphasizes the importance of considering the type of motivation behind smartphone usage when assessing its impact on health outcomes (Taylor et al., 2024). For instance, smartphones might be increasingly used to stay updated with news and maintain social connections which can have positive outcomes (Roberts and David, 2023), or more negatively used to passively monitor others’ online lives. Although this dichotomous view has been recently challenged (Valkenburg et al., 2022b), motivations behind use, and a range of other variables, can all influence the effect of screentime between users and within users over time (Orben et al., 2022; Valkenburg et al., 2022a; Vuorre et al., 2021). The present study did not assess the type of usage nor motivations for personal digital device usage, which could contribute to the null findings in the count models. However, we did explore whether steps taken per day was associated with real-world measures of anxiety, distress, and depression given the established relationship between screentime, sedentary behaviors, and cognition (Firth et al., 2024; Walsh et al., 2018; Woessner et al., 2021). Higher physical activity was associated with lower severity of depression and distress in line with previous work (Brüchle et al., 2021a; Buschert et al., 2019; Mizrahi et al., 2023). We did not observe an association between physical activity and anxiety which may be due to the type of physical activity being considered herein (i.e. steps per day). Prior work indicates that the mode and intensity of physical activity can differentially affect patient symptoms with yoga and mind-body activities having the greatest effect on anxiety relief (Singh et al., 2023). Future research should consider not only the quantity of physical activity in relation to psychiatric symptoms but also the type and intensity in relation to other behaviors such as screentime.

The present study used the D_EP_NA model to determine how each of the symptom severity and activity measures captured in the real-world influenced one another over time. This is the first application of this type of directed graph network analysis to digital phenotyping data in mood and anxiety disorders. The D_EP_NA model uniquely provides a temporally-directed measure of partial correlation effects such that a correlational influence over time can be determined. The model revealed that, rather than symptom severity per se, it was the putative underlying measures of intrinsic and extrinsic motivation that had the greatest influence over symptoms and activity in the MA group. This suggests that measures of cognitive constructs related to drive and activity may be more useful in characterizing phenotypes in the real-world. Further work should explore other cognitive measures that have been linked to mood and anxiety disorders such as executive function or sleep disturbance. In the MA group, depression was the symptom domain that was most influenced by the other measures, suggesting that this particular symptom domain is most malleable or receptive to change. This coincides with the high variability we observed of this measure over time (see **Supplementary Figure 4**). Interestingly, intrinsic motivation had amongst the greatest influence on the other measures over time and seemed to influence physical activity more in the MA group. Previous work demonstrates a link between higher physical activity and lower mood and anxiety symptoms (Brüchle et al., 2021b; Buschert et al., 2019; Hird et al., 2024). Together, this suggests that in this population, intrinsic motivation might act through physical activity to modulate symptom severity.

Recent work on symptom dynamics demonstrates that despite different patients presenting with the same level of depression severity, there are underlying differences in how symptoms are interacting with one another to drive this severity (Ebrahimi et al., 2024). The current findings also highlight the importance of assessing motivational changes in relation to mood and anxiety disorders. Indeed, prior work demonstrates that motivational deficits undermine functioning in patients with depression (Fervaha et al., 2016) and that secondary effects on specific symptoms can occur through changes in other symptoms (Bekhuis et al., 2018; Boschloo et al., 2019). Therefore, the assessment of motivation levels could 1) serve as an important risk factor for mood and anxiety disorders, 2) aid in understanding symptoms that are prone to exacerbating into a depressive episode, and 3) serve as a surrogate endpoint in clinical trials where the primary endpoint is unmet. This latter point speaks to the need to investigate the effect of treatments on individual symptoms as opposed to a summed score.

There are several limitations of this study that could be addressed in future research. First, there was missing data across EMAs that could be attributed to participant’s either skipping certain surveys or a technical issue with the application failing to administer surveys on a given day. Despite this missingness, we still observed a good level of adherence to the study and were able to maintain at least 3 days of survey data per participant. In line with this limitation, we would like to recognize our use of ZIP models as it speaks to new analytical methods that must be applied with the kind of large-scale, time-series data that comes from digital phenotyping (Reinertsen and Clifford, 2018). Indeed, these data often present challenges wherein capturing multiple measures, data points are no longer independent of one another, inter-individual variability arises, and there are often multiple variables interacting with one another.

Therefore, we call for future researchers to visually inspect their data and assess for zero-inflation when there are control groups that may inevitably report zero symptoms. Further, we suggest the use of D_EP_NA or network models to assess how different variables interact with one another given the dynamics of mood. Second, models assessing the relationship between extrinsic and intrinsic motivation on anxiety indicated residual dispersion, as did the models for screentime and those assessing the relationship between anxiety and steps (see *Supplementary Materials* for tables and figures). Therefore, while we include these results, we adhere caution with their interpretation. Third, the steps data downloaded from the *mindLAMP* server came from two different sources: 1) a pedometer and 2) Apple health. Within these data, there were instances in which the source was not clearly specified, and null values were assumed to come from the pedometer after consulting with the application’s platform developers. It is unclear whether the pedometer or Apple health has greater sensitivity, however, we did find a moderate correlation between the two sources (see **Supplementary Figure 1**). Therefore, we do not expect that the choice of data source would alter the presented results. Nonetheless, future researchers may capture greater sensitivity of movement by measuring the accelerometer data or using wearable devices. Finally, we acknowledge the lack of diversity of our study sample, which may raise concerns about the lack of representation of ethnic minorities in research studies. Although digital phenotyping aims to offer insights into real-world patient populations, its effectiveness is compromised when certain groups are excluded.

This exclusion can stem from factors such as lower smartphone ownership, digital literacy, or limited access to healthcare, which in turn exacerbates these groups’ vulnerability to mental health issues. To ensure greater access and a greater representation of ethnic diversity, we encourage researchers to supply digital devices and engage in efforts to actively recruit participants from underserved communities.

In conclusion, this study presents novel real-world measures of anxiety, distress and depression symptom severity in individuals with mood and anxiety disorders, that corresponded well to gold-standard in-lab measures. These findings highlight the potential of digital phenotyping for accurately assessing and monitoring psychiatric conditions with good adherence. Furthermore, using a combination of ZIP models and network analysis, the presented work highlights how underlying cognitive measures such as intrinsic and extrinsic motivation may be most influential in predicting symptom severity and physical/digital activity.

## Acknowledgements

We would like to thank all the participants who took part in this study and The Charles Bronfman Institute for Personalized Medicine for their technological resources, maintenance, and staff expertise. Illustrations in Figure 1 were created with BioRender.com. Funding was provided by NIMH K01MH12043, the Friedman Brain Institute, the Ehrenkranz Laboratory for Human Resilience, and the Gottesman Foundation. This work was supported in part through the computational and data resources and staff expertise provided by Scientific Computing and Data at the Icahn School of Medicine at Mount Sinai and supported by the Clinical and Translational Science Awards (CTSA) grant UL1TR004419 from the National Center for Advancing Translational Sciences.

## Data Availability

The data that support the findings of the current study are available from the corresponding author upon reasonable request.

## Code Availability

The underlying code for this study is not publicly available but may be made available to qualified researchers on reasonable request from the corresponding author.

