## Supplementary Materials for "Relationships between depression, anxiety, and motivation in the real-world: Effects of physical activity and screentime" (from another version of this preprint: version 1, 2024-08-07)

### Supplementary Material

#### Supplementary Results

**Result 1.** Relationships between real-world and in-lab measures of symptom severity

*Anxiety*

In a model with the full cohort, a 1-point increase in the MASQ Anxious Arousal score was associated with a 0.1-point increase in real-world anxiety, adjusting for day and group (see **Figure 4** in main text). Within each cohort the association differed wherein the coefficient was 2.22-times larger in the HC ($\hat{\beta} = 0.16$) compared to the MA group ($\hat{\beta} = 0.07$). Specifically, the MA group had a 0.07-point increase in real-world anxiety for every one-point increase in the MASQ Anxious Arousal score adjusting for day and group; HC had a 0.16-point increase. In the full cohort model, those in the MA group were 1.55-points higher on the real-world scale relative to those in the HC group (t = 3.45, p = 0.0006). Within the anxious real-world scale, the full cohort model revealed an ICC of 0.70, indicating moderate reliability (**Supplementary Figure 5**). Within each cohort, the HC group had good-to-excellent reliability (ICC = 0.76), whereas the MA group had moderate reliability (ICC = 0.58) (**Supplementary Figure 5).**

*Distress*

In a model with the full cohort, a 1-point increase in the MASQ General Distress score was associated with a 0.14-point increase in real-world distress, adjusting for day and group. Within each cohort the association differed wherein the coefficient was roughly 2.7-times larger in the HC group ($\hat{\beta} = 0.30$) compared to the MA group ($\hat{\beta} = 0.11$). Specifically, the MA group had a 0.11-point increase in real-world distress for every one-point increase in the MASQ General Distress score adjusted for day and group; HC had a 0.30-point increase. In the full cohort model, those in the MA group were 1.60-points higher on the real-world scale relative to those in the HC group (t = 3.93, p < 0.001). Within the total score for the real-world scale, the full cohort model revealed an ICC of 0.67, indicating moderate reliability (**Supplementary Figure 5**). When stratified by cohort, the ICC for the HC group (ICC = 0.74) indicated moderate reliability, however, the MA group had poor reliability (ICC = 0.49) (**Supplementary Figure 5**).

*Depression*

In a model with the full cohort, a 1-point increase in the MASQ Anhedonic Depression score was associated with a 0.08-point increase in real-world depression, adjusting for day and group. Within each cohort the association differed wherein the coefficient was roughly 2-times larger in the MA group ($\hat{\beta} = 0.11$) compared to the HC group ($\hat{\beta} = 0.05$). Specifically, the MA group had a 0.11-point increase in real-world depression for every one-point increase in the MASQ Anhedonic Depression score adjusted for day; HC had a 0.05-point increase. In the full cohort model, those in the MA group were 2.43-points higher on the real-world scale relative to those in the HC group (t = 4.9, p < 0.001). Within the depression real-world scale, the full cohort model revealed an ICC of 0.75, indicating good-to-excellent reliability **(Supplementary Figure 5**). Both groups showed moderate reliability, with the HC group (ICC = 0.68) having a higher ICC than the MA group (ICC = 0.63) (**Supplementary Figure 5**).

*Total score*

In the full cohort, a 1-point increase in the MASQ Total Score was associated with a 0.20-point increase in the real-world anxiety/distress/depression total score, adjusting for day and group. Within each cohort the association differed wherein the coefficient was roughly 1.4-times larger in the MA group ($\hat{\beta} = 0.23$) compared to the HC group ($\hat{\beta} = 0.17$). Specifically, the MA group had a 0.23-point increase on the real-world scale for every one-point increase in the MASQ Total Score adjusted for day; HC had a 0.17-point increase. In the full cohort model, those in the MA group were 2.84-points higher on the real-world scale relative to those in the HC group ($t = 2.45$, $p = 0.014$). Within the total score for the real-world scale, the full cohort model revealed an ICC of 0.78, indicating good-to-excellent reliability, as did the HC group (ICC = 0.78), but not the MA group (ICC = 0.64) which had moderate reliability **(Supplementary Figure 5)**.

### Supplementary tables and figures

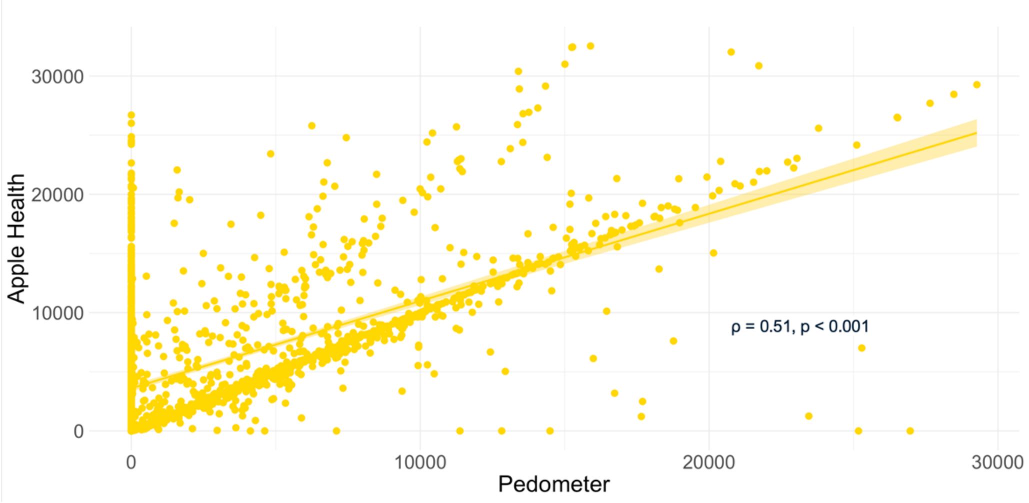


**Supplementary Figure 1.** The correlation between steps data captured by the pedometer and Apple Health was moderate (N=73, rho=0.51, $p < 0.001$). Note, one datapoint with pedometer value = 71,132 and Apple Health value = 18,963 was omitted. Including this datapoint did not impact the results (N=73, rho=0.51, $p < 0.001$).

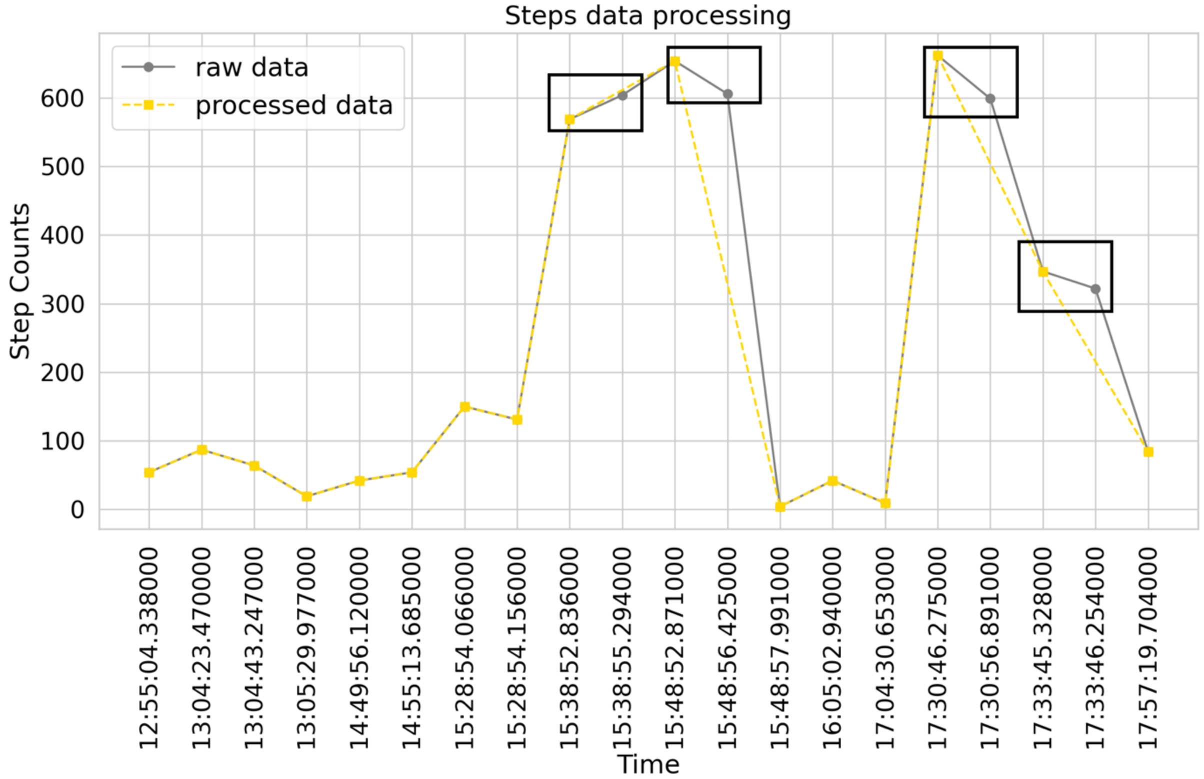


**Supplementary Figure 2.** Example 20 timepoints for one participant's step counts data before (raw data) and after preprocessing (processed data). Values recorded within 30 seconds of each other and within 10% magnitude of each other were considered duplicates and only the first entry was included to account for a sensor-related error-margin. Black boxes denote timepoints considered as duplicates based on the aforementioned criteria.

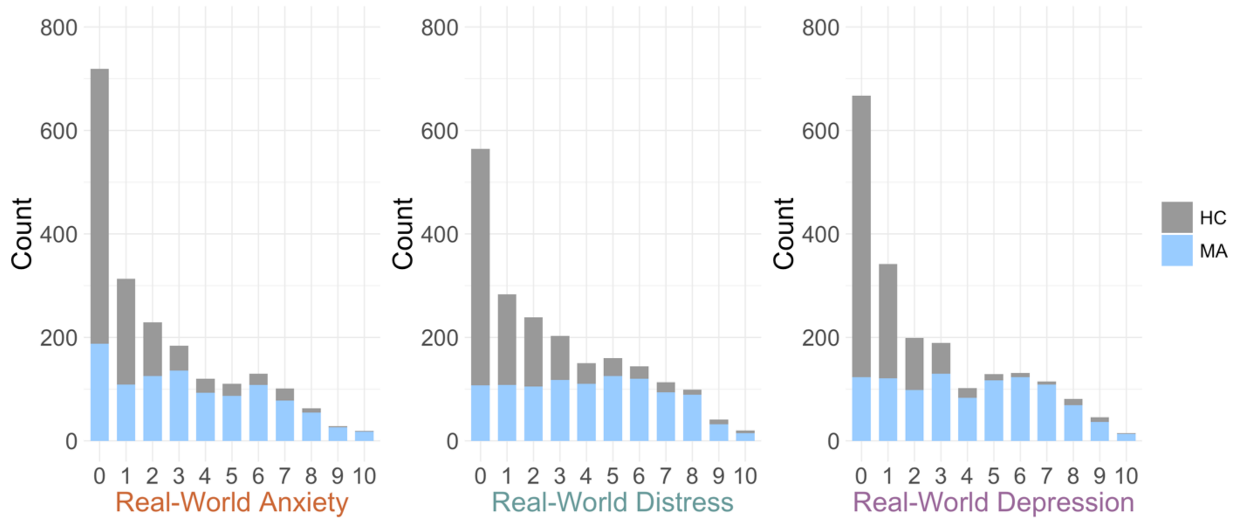


**Supplementary Figure 3.** Distribution of aggregated scores for real-world anxiety, distress, and depression between groups. Here, we gauge that most HCs report zero symptoms resulting in notable zero-inflation in our data. *HC = Healthy Control group, MA = Mood/Anxiety disorder group*

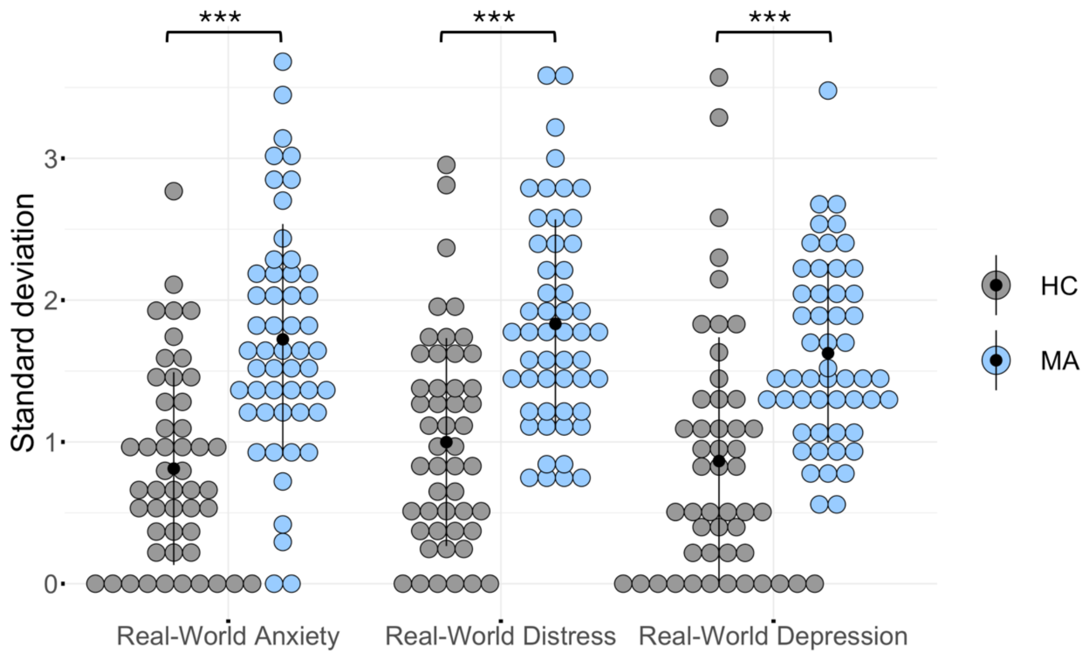


**Supplementary Figure 4.** Standard deviation of real-world anxiety/distress/depression symptoms reported over time between groups. The variability of the three real-world measures was higher in the MA group compared to HC group across all three measures (Real-World Anxiety/Distress/Depression: $t_{(99)} = -6.09$, $p < 0.001$, $t_{(99)} = -5.70$, $p < 0.001$, $t_{(87.1)} = -4.99$, $p < 0.001$)

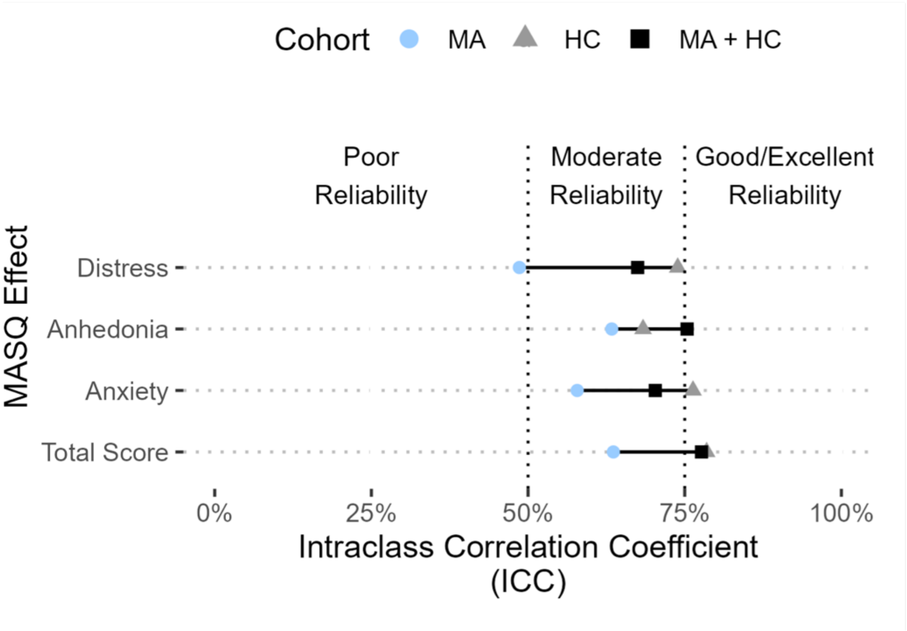


**<u>Supplementary Figure 5.</u>** Assessment of the internal consistency of the real-world scales over time. Values below 0.5 indicate poor reliability, between 0.5 and 0.75 indicate moderate reliability, and any value above 0.75 indicates good-to-excellent reliability (Koo and Li, 2016).

**Relationships between real-world and in-lab measures of symptom severity**

**Table 1.** Association between real-world anxiety and the MASQ Anxious Arousal (AA) scale

| | Real-World Anxiety ~ MASQ AA + day + (1\|participant) | |
|---|---|---|
| *Predictors* | *Estimates* | *p* |
| (Intercept) | 1.80 (0.15 – 3.45) | **0.038** |
| MASQ AA | 0.07 (0.01 – 0.13) | **0.024** |
| day | 0.007 (-0.01 – 0.02) | 0.304 |
| **Random Effects** | | |
| $\sigma^2$ | 3.16 | |
| $\tau_{00\ participant}$ | 3.78 | |
| ICC | 0.54 | |
| $N_{participant}$ | 47 | |
| Observations | 957 | |
| Marginal $R^2$/Conditional $R^2$ | 0.060 / 0.572 | |

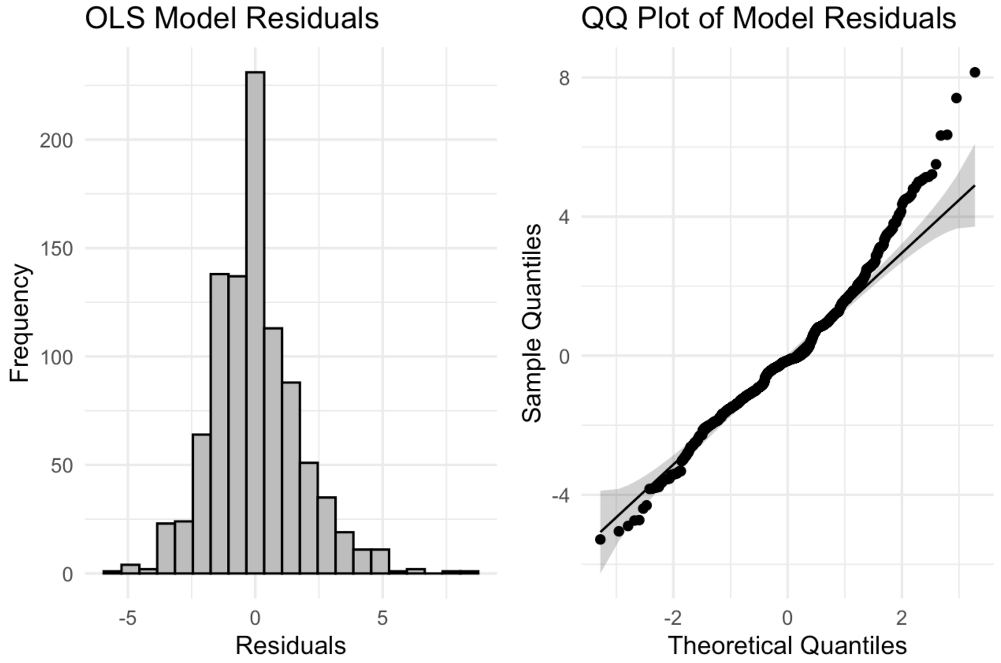

**Table 2.** Association between real-world distress and the MASQ General Distress (GD) scale

| | Real-World Distress ~ MASQ GD + day + (1\|participant) | |
|---|---|---|
| *Predictors* | *Estimates* | *p* |
| (Intercept) | 2.18<br>(1.03 – 3.33) | **<0.001** |
| MASQ GD | 0.11<br>(0.06 – 0.16) | **<0.001** |
| day | -0.01<br>(-0.02 – 0.00) | 0.169 |
| **Random Effects** | | |
| $\sigma^2$ | 3.70 | |
| $\tau_{00\ participant}$ | 2.26 | |
| ICC | 0.38 | |
| $N_{participant}$ | 47 | |
| Observations | 957 | |
| Marginal $R^2$/Conditional $R^2$ | 0.164 / 0.481 | |

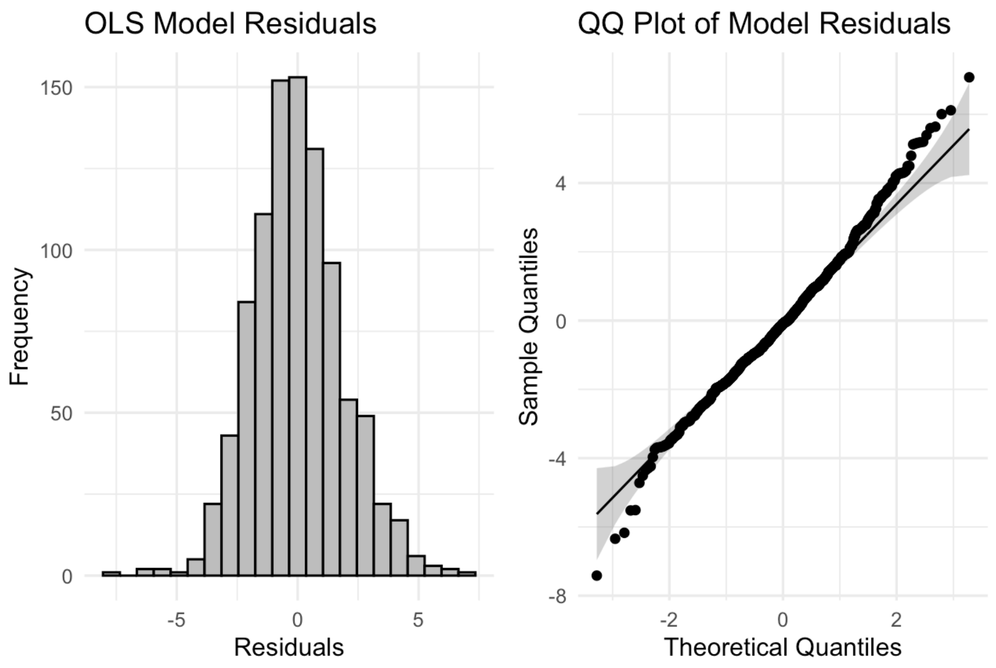

**Table 3.** Association between real-world depression and the MASQ Anhedonic Depression (AD) scale

| | Real-World Depression ~ MASQ AD + day + (1\|participant) | |
|---|---|---|
| *Predictors* | *Estimates* | *p* |
| (Intercept) | 0.19<br>(-2.95 – 3.33) | 0.905 |
| MASQ AD | 0.11<br>(0.03 – 0.19) | **0.006** |
| day | -0.01<br>(-0.02 – 0.00) | 0.097 |
| **Random Effects** | | |
| $\sigma^2$ | 2.72 | |
| $\tau_{00\ participant}$ | 3.94 | |
| ICC | 0.59 | |
| $N_{participant}$ | 47 | |
| Observations | 949 | |
| Marginal $R^2$/Conditional $R^2$ | 0.095 / 0.630 | |

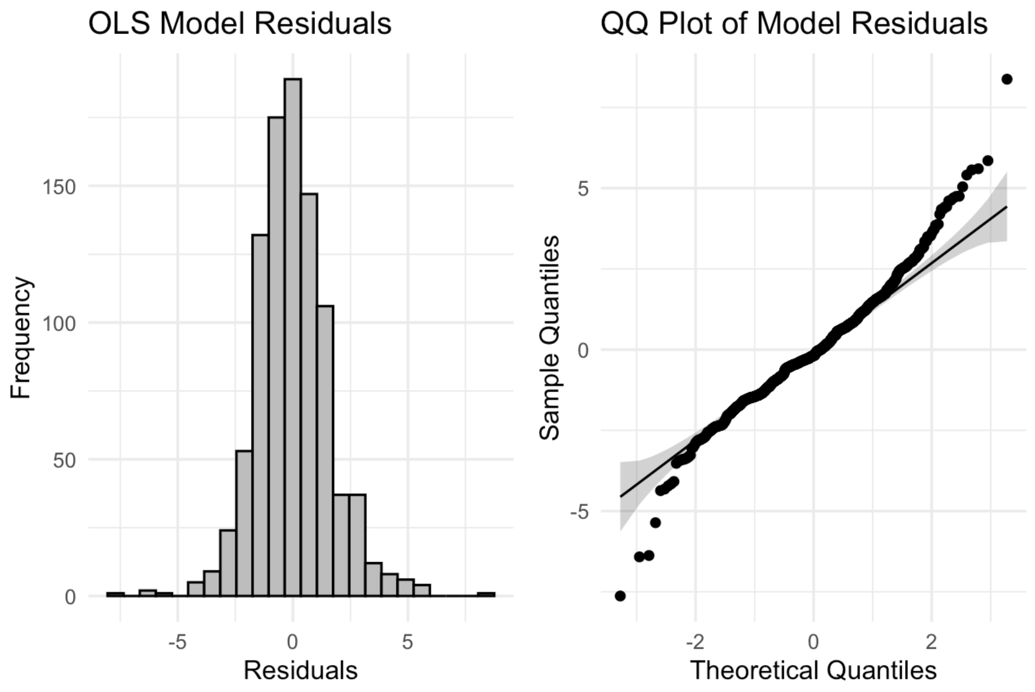

**Exploring the effects of intrinsic and extrinsic motivation on anxiety, distress, and depression**

**Table 4.** Association between real-world anxiety and intrinsic motivation

| | Real-World Anxiety ~ Intrinsic Motivation*group + day + (1\|participant)<br>Logit: ~ Intrinsic Motivation + group + (1\|participant) | |
|---|---|---|
| *Predictors* | *Incidence Rate Ratios* | *p* |
| **Count Model** | | |
| (Intercept) | 3.33<br>(2.32 – 4.78) | **<0.001** |
| Intrinsic motivation | 0.87<br>(0.83 – 0.90) | **<0.001** |
| Group [MA] | 1.23<br>(0.83 – 1.83) | 0.303 |
| day | 1.00<br>(1.00 – 1.00) | 0.447 |
| Intrinsic motivation x group [MA] | 1.12<br>(1.07 – 1.17) | **<0.001** |
| **Logit Model** | | |
| **(Intercept)** | 0.002<br>(0.00 – 0.02) | **<0.001** |
| Intrinsic motivation | 1.62<br>(1.29 – 2.04) | **<0.001** |
| Group [MA] | 0.20<br>(0.03 – 1.58) | 0.127 |
| **Random Effects** | | |
| $\sigma^2$ | 0.35 | |
| $\tau_{00\ participant}$ | 0.31 | |
| ICC | 0.47 | |
| $N_{participant}$ | 101 | |
| Observations | 2072 | |
| Marginal $R^2$/Conditional $R^2$ | 0.374/ 0.668 | |

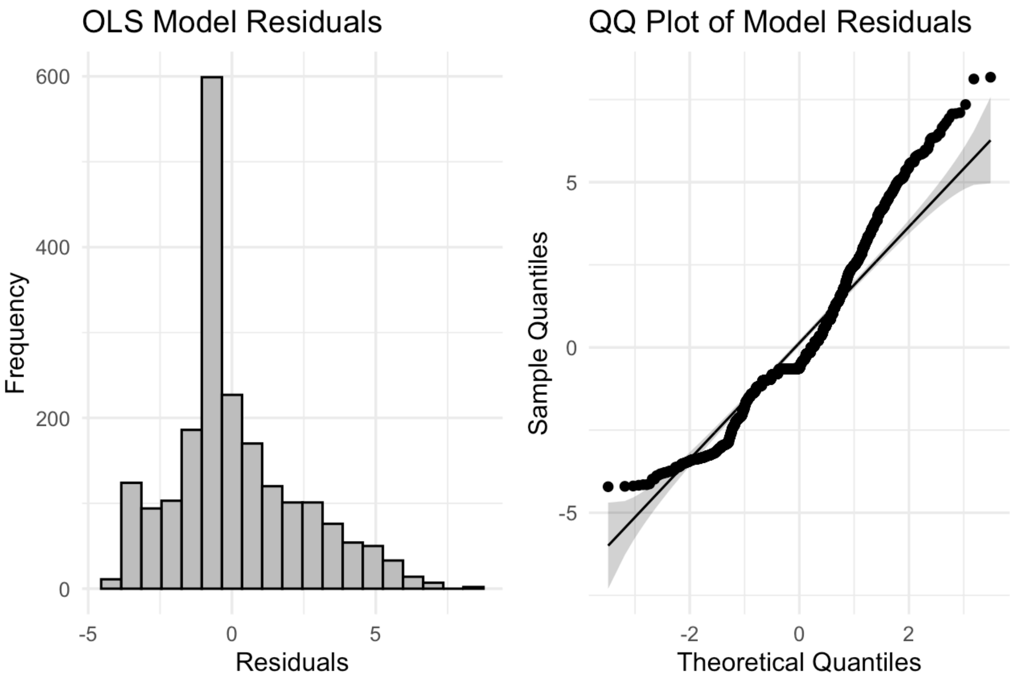

OLS Model Residuals
Frequency
600
400
200
0
-5
0
5
Residuals
QQ Plot of Model Residuals
Sample Quantiles
5
0
-5
-2
0
2
Theoretical Quantiles

**Table 5.** Association between real-world distress and intrinsic motivation

| *Predictors* | Real-World Distress ~ Intrinsic Motivation*group + day + (1\|participant)<br>Logit: ~ Intrinsic Motivation + day + group + (1\|participant)<br>*Incidence Rate Ratios* | *p* |
|---|---|---|
| **Count Model** | | |
| (Intercept) | 5.15<br>(3.81 – 6.96) | **<0.001** |
| Intrinsic motivation | 0.86<br>(0.83 – 0.89) | **<0.001** |
| Group [MA] | 1.21<br>(0.87 – 1.69) | 0.260 |
| day | 1.00<br>(1.00 – 1.00) | 0.941 |
| Intrinsic motivation x group [MA] | 1.08<br>(1.04 – 1.12) | **<0.001** |
| **Logit Model** | | |
| **(Intercept)** | 0.00<br>(0.00 – 0.00) | **<0.001** |
| Intrinsic motivation | 2.67<br>(1.85 – 3.85) | **<0.001** |
| Day | 1.06<br>(1.02 – 1.10) | **0.003** |
| Group [MA] | 0.11<br>(0.02 – 0.69) | **0.019** |
| **Random Effects** | | |
| $N_{participant}$ | 101 | |
| Observations | 2076 | |

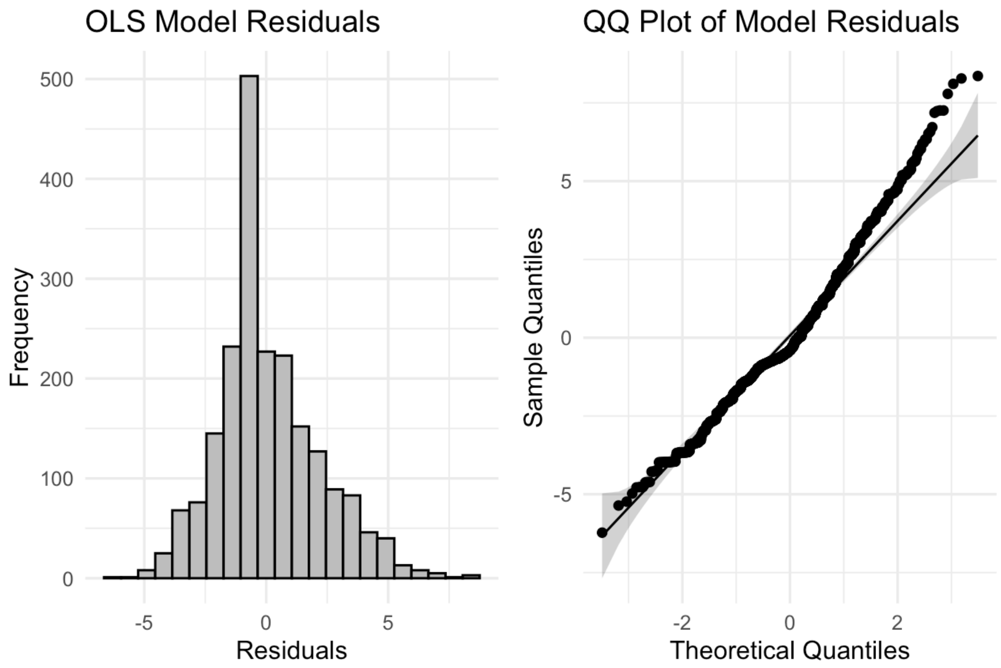
OLS Model Residuals
Frequency
500
400
300
200
100
0
-5
0
5
Residuals
QQ Plot of Model Residuals
Sample Quantiles
5
0
-5
-2
0
2
Theoretical Quantiles

**Table 6.** Association between real-world depression and intrinsic motivation

| *Predictors* | Real-World Depression ~ Intrinsic Motivation*group + day + (1\|participant)<br>Logit: ~ Intrinsic Motivation + day + group + (1\|participant)<br>*Incidence Rate Ratios* | *p* |
|---|---|---|
| **Count Model** | | |
| (Intercept) | 5.83<br>(4.15 – 8.18) | **<0.001** |
| Intrinsic motivation | 0.82<br>(0.79 – 0.86) | **<0.001** |
| Group [MA] | 1.10<br>(0.76 – 1.58) | 0.629 |
| day | 1.00<br>(1.00 – 1.00) | 0.886 |
| Intrinsic motivation x group [MA] | 1.11<br>(1.06 – 1.17) | **<0.001** |
| **Logit Model** | | |
| **(Intercept)** | 0.00<br>(0.00 – 0.00) | **<0.001** |
| Intrinsic motivation | 2.29<br>(1.60 – 3.29) | **<0.001** |
| Day | 1.05<br>(1.01 – 1.10) | **0.013** |
| Group [MA] | 0.01<br>(0.00 – 0.16) | **<0.001** |
| **Random Effects** | | |
| $\sigma^2$ | 0.28 | |
| $\tau_{00\ participant}$ | 0.21 | |
| ICC | 0.43 | |
| $N_{participant}$ | 101 | |
| Observations | 2059 | |
| Marginal $R^2$/Conditional $R^2$ | 0.478/ 0.703 | |

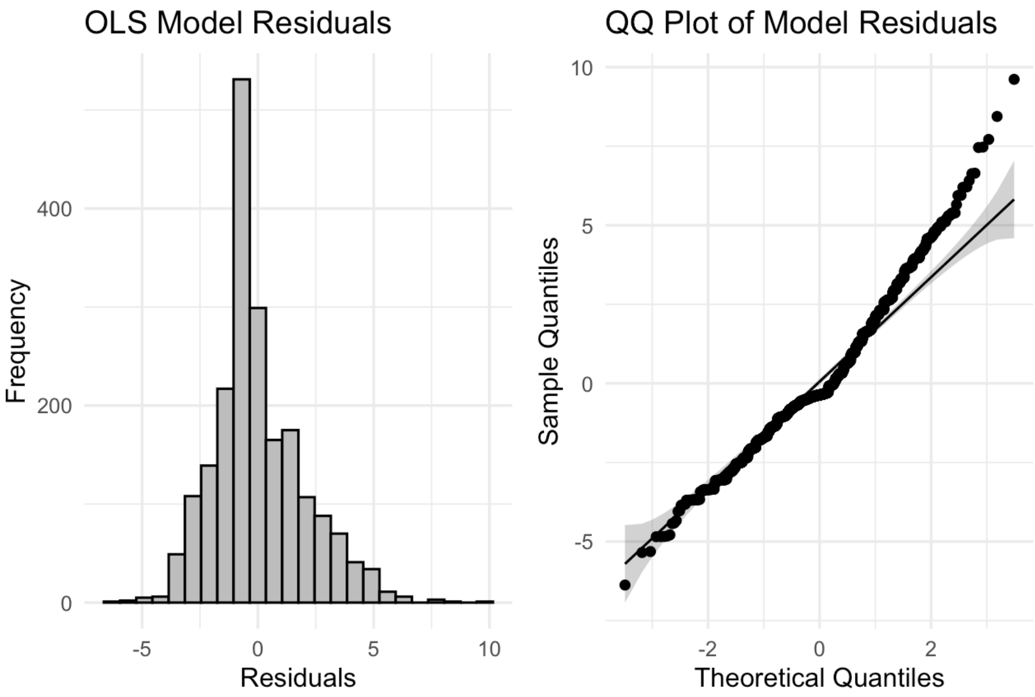
OLS Model Residuals
Frequency
400
200
0
-5
0
5
10
Residuals
QQ Plot of Model Residuals
Sample Quantiles
10
5
0
-5
-2
0
2
Theoretical Quantiles

**Table 7.** Association between real-world anxiety and extrinsic motivation

| | Real-World Anxiety ~ Extrinsic Motivation *group + day + (1\|participant)<br>Logit: ~ Extrinsic Motivation + group + (1\|participant) | |
|---|---|---|
| *Predictors* | *Incidence Rate Ratios* | *p* |
| **Count Model** | | |
| (Intercept) | 1.87<br>(1.31 – 2.68) | **0.011** |
| Extrinsic motivation | 0.93<br>(0.90 – 0.97) | **0.001** |
| Group [MA] | 2.07<br>(1.39 – 3.08) | **<0.001** |
| day | 1.00<br>(1.00 – 1.00) | 0.676 |
| Extrinsic motivation x group [MA] | 1.05<br>(1.01 – 1.10) | **0.024** |
| **Logit Model** | | |
| **(Intercept)** | 0.00<br>(0.00 – 0.04) | **<0.001** |
| Extrinsic motivation | 1.55<br>(1.23 – 1.97) | **<0.001** |
| Group [MA] | 0.15<br>(0.02 – 1.23) | 0.077 |
| **Random Effects** | | |
| $\sigma^2$ | 0.36 | |
| $\tau_{00\ participant}$ | 0.39 | |
| ICC | 0.52 | |
| $N_{participant}$ | 101 | |
| Observations | 2060 | |
| Marginal $R^2$/Conditional $R^2$ | 0.317/ 0.669 | |

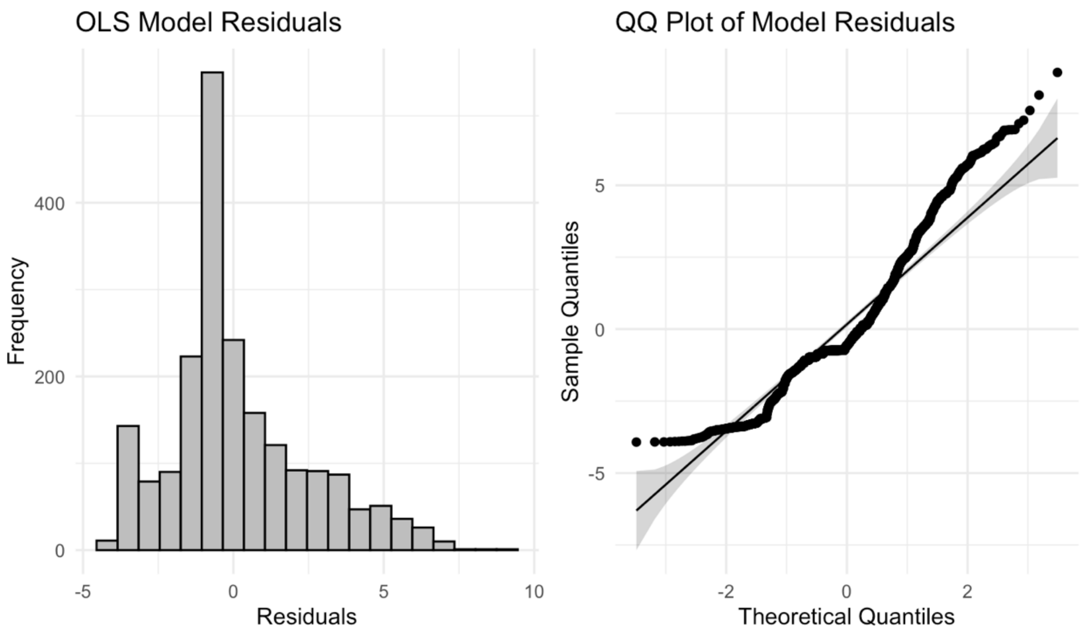
OLS Model Residuals
Frequency
400
200
0
-5
0
5
10
Residuals
QQ Plot of Model Residuals
Sample Quantiles
5
0
-5
-2
0
2
Theoretical Quantiles

**Table 8.** Association between real-world distress and extrinsic motivation

| | Real-World Distress ~ Extrinsic Motivation*group + day +(1\|participant)<br>Logit: ~ Extrinsic Motivation + day + group + (1\|participant) | |
|---|---|---|
| *Predictors* | *Incidence Rate Ratios* | *p* |
| **Count Model** | | |
| (Intercept) | 2.70<br>(1.98 – 3.68) | **<0.001** |
| Extrinsic motivation | 0.93<br>(0.90 – 0.97) | **<0.001** |
| Group [MA] | 2.02<br>(1.42 – 2.87) | **<0.001** |
| day | 1.00<br>(1.00 – 1.00) | 0.431 |
| Extrinsic motivation x group [MA] | 1.02<br>(0.98 – 1.06) | 0.297 |
| **Logit Model** | | |
| **(Intercept)** | 0.00<br>(0.00 – 0.00) | **<0.001** |
| Extrinsic motivation | 2.22<br>(1.54 – 3.19) | **<0.001** |
| Day | 1.05<br>(1.01 – 1.09) | **0.009** |
| Group [MA] | 0.09<br>(0.01 – 0.57) | **0.010** |
| **Random Effects** | | |
| $\sigma^2$ | 0.31 | |
| $\tau_{00\ participant}$ | 0.31 | |
| ICC | 0.50 | |
| $N_{participant}$ | 101 | |
| Observations | 2062 | |
| Marginal $R^2$/Conditional $R^2$ | 0.291/ 0.644 | |

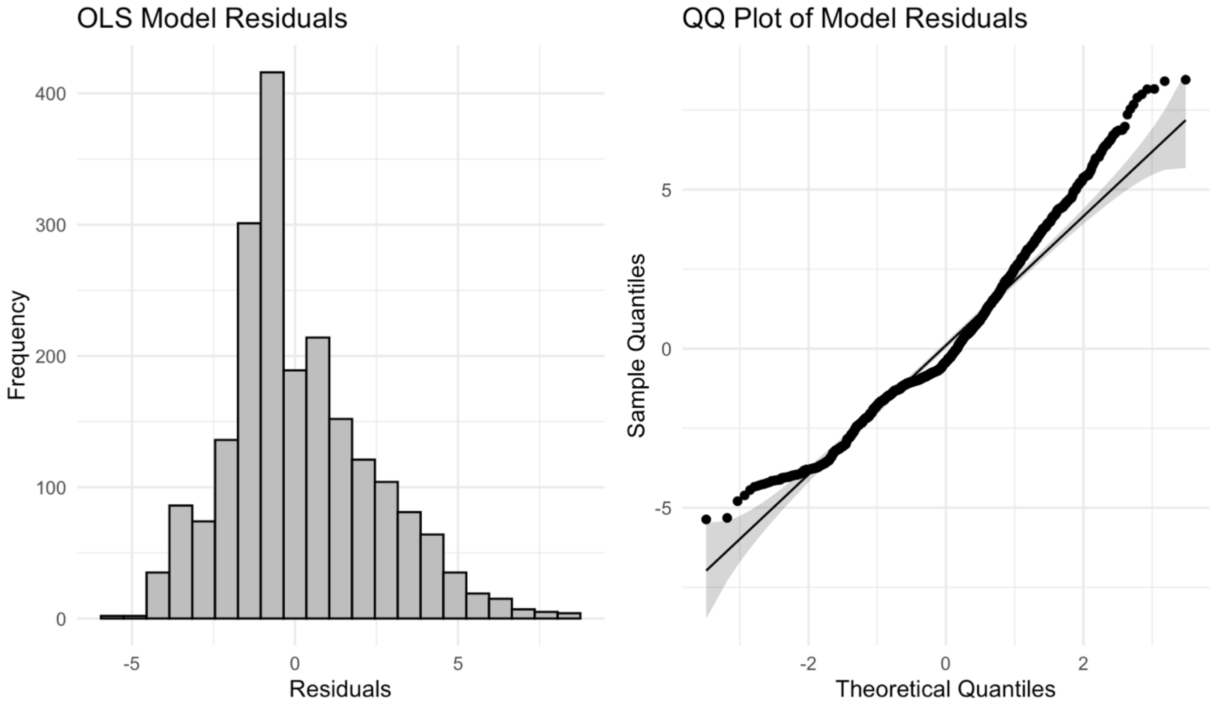
OLS Model Residuals
Frequency
Residuals
400
300
200
100
0
-5
0
5
QQ Plot of Model Residuals
Sample Quantiles
Theoretical Quantiles
5
0
-5
-2
0
2

**Table 9.** Association between real-world depression and extrinsic motivation

| | Real-World Depression ~ Extrinsic Motivation*group + day +(1\|participant)<br>Logit: ~ Extrinsic Motivation + group + (1\|participant) | |
|---|---|---|
| *Predictors* | *Incidence Rate Ratios* | *p* |
| **Count Model** | | |
| (Intercept) | 3.54<br>(2.50 – 5.01) | **<0.001** |
| Extrinsic motivation | 0.88<br>(0.85 – 0.92) | **<0.001** |
| Group [MA] | 1.70<br>(1.16 – 2.48) | **0.007** |
| day | 1.00<br>(0.99 – 1.00) | **0.059** |
| Extrinsic motivation x group [MA] | 1.06<br>(1.01 – 1.10) | **0.019** |
| **Logit Model** | | |
| **(Intercept)** | 0.02<br>(0.00 – 0.24) | **0.002** |
| Extrinsic motivation | 1.33<br>(1.06 – 1.65) | **0.012** |
| Group [MA] | 0.01<br>(0.00 – 0.06) | **<0.001** |
| **Random Effects** | | |
| $\sigma^2$ | 0.29 | |
| $\tau_{00\ participant}$ | 0.26 | |
| ICC | 0.47 | |
| $N_{participant}$ | 101 | |
| Observations | 2046 | |
| Marginal $R^2$/Conditional $R^2$ | 0.396 / 0.682 | |

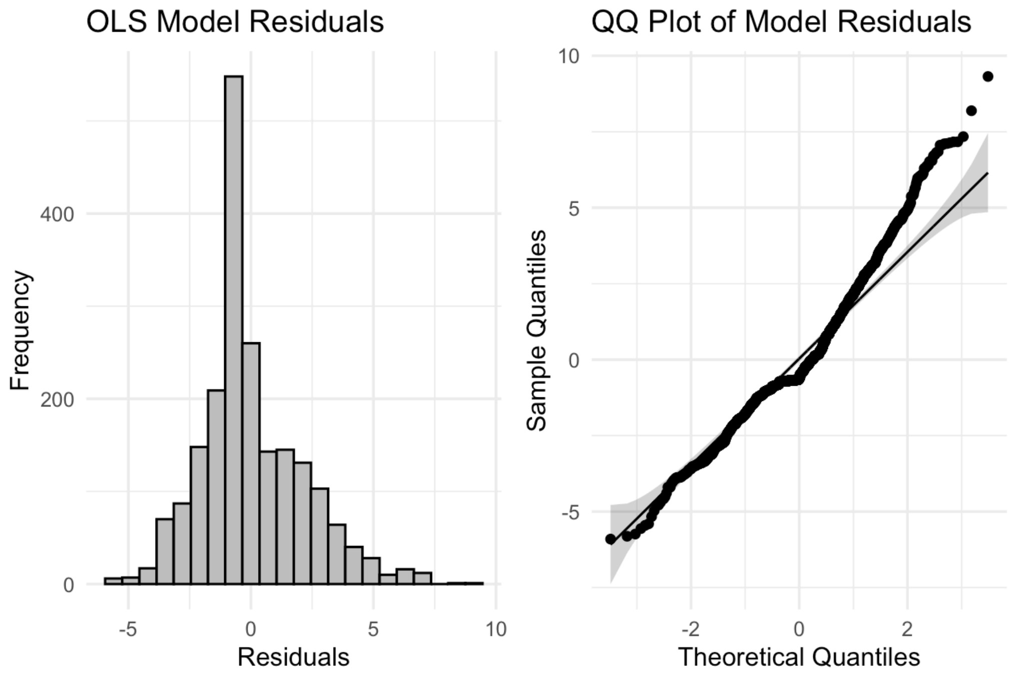

OLS Model Residuals
Frequency
400
200
0
-5
0
5
10
Residuals
QQ Plot of Model Residuals
Sample Quantiles
10
5
0
-5
-2
0
2
Theoretical Quantiles

**Exploring the effects of activity on anxiety, distress and depression**

**Table 10.** Association between real-world anxiety and steps

| | Real-World Anxiety ~ Steps + day + group + (1\|participant)<br>Logit: ~ steps + day + group + (1\|participant) | |
|---|---|---|
| *Predictors* | *Incidence Rate Ratios* | *p* |
| **Count Model** | | |
| (Intercept) | 1.23<br>(0.90 – 1.69) | 0.194 |
| Steps | 0.98<br>(0.94 – 1.03) | 0.461 |
| Group [MA] | 3.28<br>(2.29 – 4.69) | **< 0.001** |
| Day | 1.00<br>(1.00 – 1.00) | 0.939 |
| **Logit Model** | | |
| **(Intercept)** | 0.22<br>(0.02 – 2.96) | 0.256 |
| Steps | 1.09<br>(0.69 – 1.73) | 0.716 |
| Day | 0.98<br>(0.93 – 1.03) | 0.478 |
| Group [MA] | 0.06<br>(0.00 – 1.03) | **0.052** |
| **Random Effects** | | |
| $\sigma^2$ | 0.33 | |
| $\tau_{00\ participant}$ | 0.35 | |
| ICC | 0.51 | |
| $N_{participant}$ | 70 | |
| Observations | 1235 | |
| Marginal $R^2$/Conditional $R^2$ | 0.344/ 0.681 | |

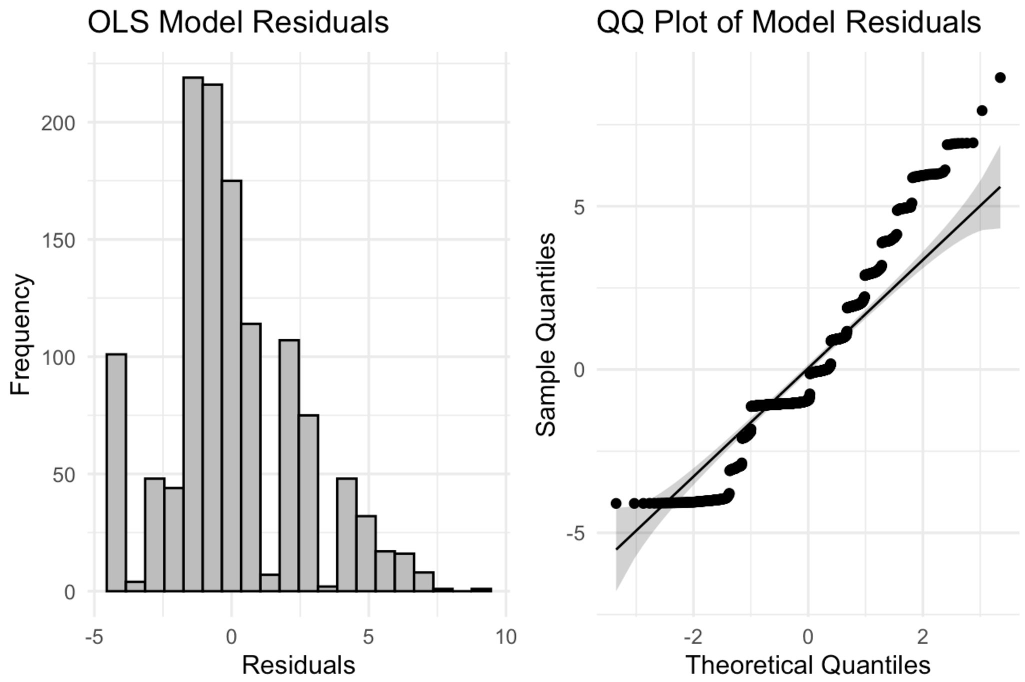

OLS Model Residuals
Frequency
Residuals
0
50
100
150
200
-5
0
5
10
QQ Plot of Model Residuals
Sample Quantiles
Theoretical Quantiles
-5
0
5
-2
0
2

**Table 11.** Association between real-world distress and steps

| | Real-World Distress ~ Steps + day + group + (1\|participant)<br>Logit: ~ steps + day + group + (1\|participant) | |
|---|---|---|
| *Predictors* | *Incidence Rate Ratios* | *p* |
| **Count Model** | | |
| (Intercept) | 1.94<br>(1.52 – 2.47) | **<0.001** |
| Steps | 0.96<br>(0.91 – 1.00) | **0.059** |
| Group [MA] | 2.18<br>(1.64 – 2.92) | **< 0.001** |
| Day | 1.00<br>(0.99 – 1.00) | 0.406 |
| **Logit Model** | | |
| **(Intercept)** | 0.13<br>(0.03 – 0.59) | **0.009** |
| Steps | 1.14<br>(0.73 – 1.80) | 0.556 |
| Day | 1.05<br>(1.01 – 1.09) | **0.022** |
| Group [MA] | 0.02<br>(0.00 – 0.13) | **<0.001** |
| **Random Effects** | | |
| $\sigma^2$ | 0.27 | |
| $\tau_{00\ participant}$ | 0.26 | |
| ICC | 0.49 | |
| $N_{participant}$ | 70 | |
| Observations | 1236 | |
| Marginal $R^2$/Conditional $R^2$ | 0.232/ 0.608 | |

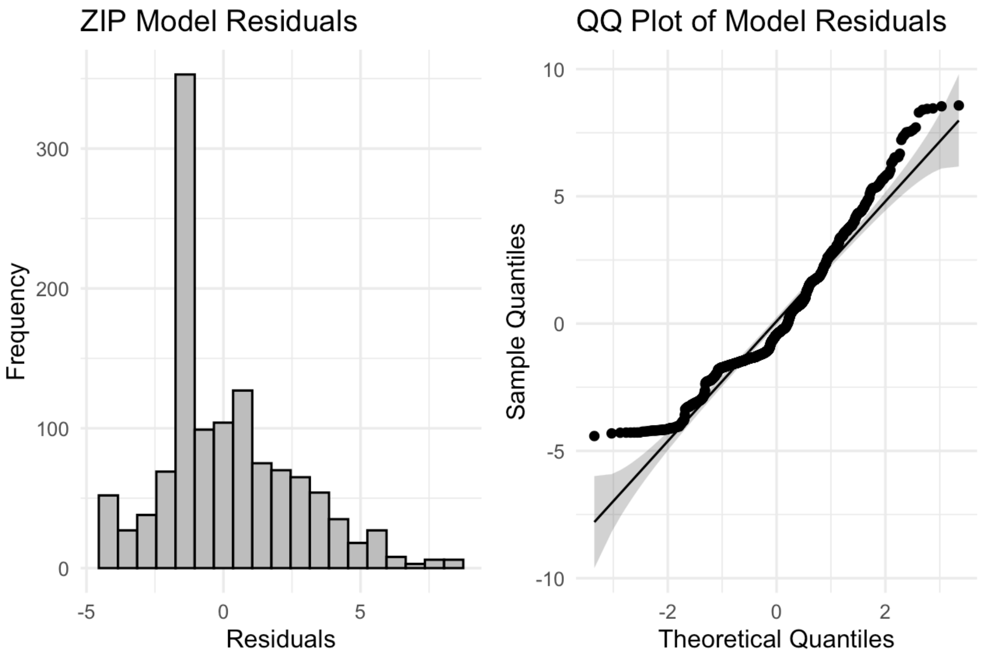

ZIP Model Residuals
Frequency
300
200
100
0
-5
0
5
Residuals
QQ Plot of Model Residuals
Sample Quantiles
10
5
0
-5
-10
-2
0
2
Theoretical Quantiles

**Table 12.** Association between real-world depression and steps

| | Real-World Depression ~ Steps + day + group + (1\|participant)<br>Logit: ~ group + (1\|participant) | |
|---|---|---|
| *Predictors* | *Incidence Rate Ratios* | *p* |
| **Count Model** | | |
| (Intercept) | 2.06<br>(1.55 – 2.73) | **<0.001** |
| Steps | 0.93<br>(0.89 – 0.98) | **0.003** |
| Group [MA] | 2.13<br>(1.54 – 2.95) | **< 0.001** |
| Day | 0.99<br>(0.99 – 1.00) | **0.025** |
| **Logit Model** | | |
| **(Intercept)** | 0.50<br>(0.06 – 4.04) | 0.518 |
| Group [MA] | 0.00<br>(0.00 – 0.01) | **<0.001** |
| **Random Effects** | | |
| $\sigma^2$ | 0.22 | |
| $\tau_{00\ participant}$ | 0.25 | |
| ICC | 0.53 | |
| $N_{participant}$ | 70 | |
| Observations | 1228 | |
| Marginal $R^2$/Conditional $R^2$ | 0.250/ 0.646 | |

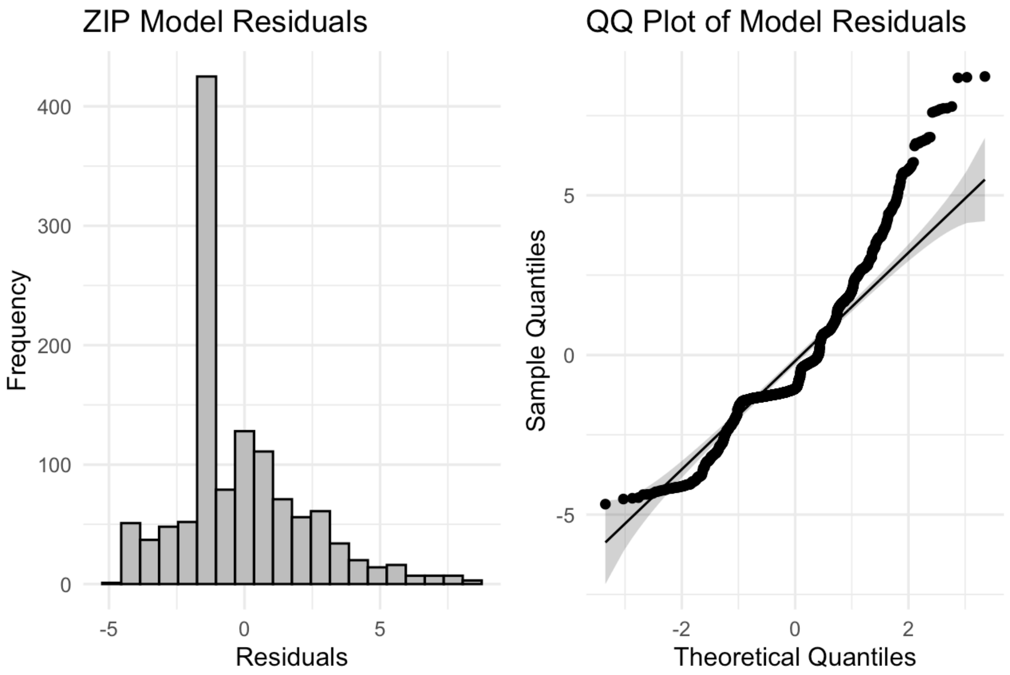
ZIP Model Residuals
Frequency
400
300
200
100
0
-5
0
5
Residuals
QQ Plot of Model Residuals
Sample Quantiles
5
0
-5
-2
0
2
Theoretical Quantiles

**Table 13.** Association between real-world anxiety and screentime

| | Real-World Anxiety ~ Screentime + day + group + (1\|participant)<br>Logit: ~ screentime + day + group + (1\|participant) | |
|---|---|---|
| *Predictors* | *Incidence Rate Ratios* | *p* |
| **Count Model** | | |
| (Intercept) | 1.20<br>(0.89 – 1.61) | 0.233 |
| Screentime | 1.02<br>(0.96 – 1.07) | 0.573 |
| Group [MA] | 2.67<br>(1.90 – 3.78) | **< 0.001** |
| Day | 1.01<br>(1.00 – 1.01) | **0.032** |
| **Logit Model** | | |
| **(Intercept)** | 0.08<br>(0.01 – 0.90) | **0.040** |
| Screentime | 1.50<br>(0.90 – 2.52) | 0.120 |
| Day | 1.04<br>(0.99 – 1.09) | 0.087 |
| Group [MA] | 0.05<br>(0.00 – 1.65) | 0.093 |
| **Random Effects** | | |
| $\sigma^2$ | 0.31 | |
| $\tau_{00\ participant}$ | 0.39 | |
| ICC | 0.56 | |
| $N_{participant}$ | 80 | |
| Observations | 1152 | |
| Marginal $R^2$/Conditional $R^2$ | 0.248/ 0.666 | |

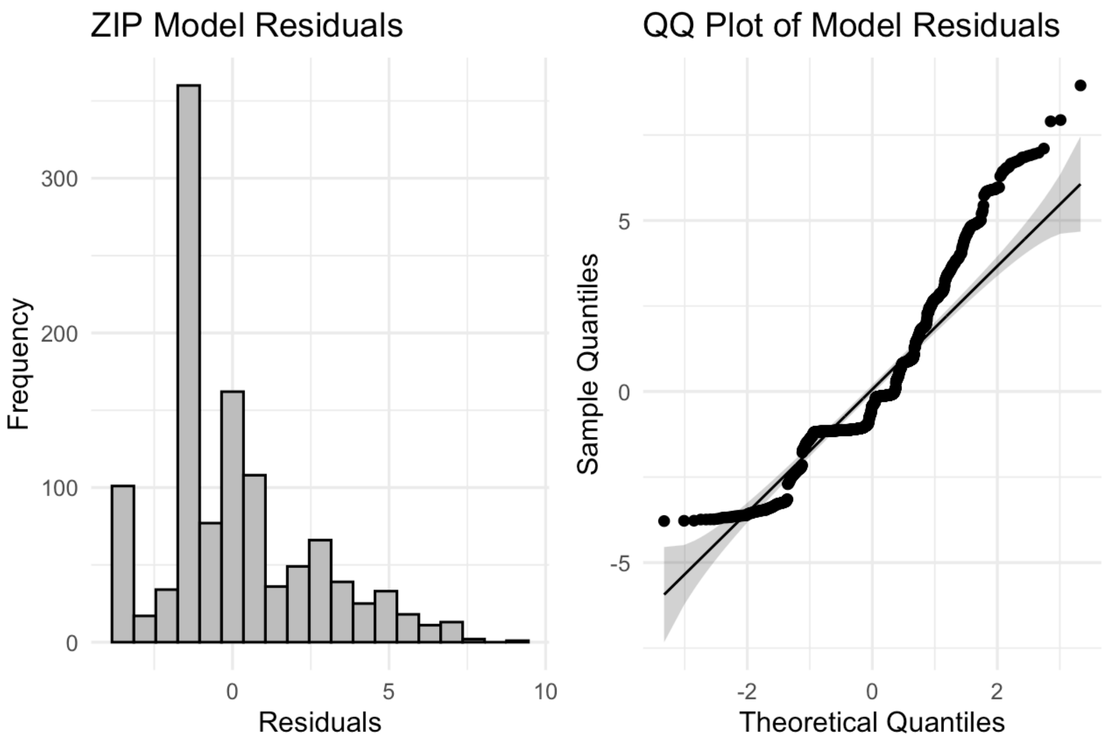

ZIP Model Residuals
Frequency
300
200
100
0
0
5
10
Residuals
QQ Plot of Model Residuals
Sample Quantiles
5
0
-5
-2
0
2
Theoretical Quantiles

**Table 14.** Association between real-world anxiety and distress

| *Predictors* | Real-World Distress ~ Screentime + day + group + (1\|participant)<br>Logit: ~ screentime + day + group + (1\|participant)<br>*Incidence Rate Ratios* | *p* |
|---|---|---|
| **Count Model** | | |
| (Intercept) | 1.79<br>(1.42 – 2.26) | **<0.001** |
| Screentime | 1.00<br>(0.95 – 1.05) | 0.987 |
| Group [MA] | 2.16<br>(1.63 – 2.86) | **< 0.001** |
| Day | 1.00<br>(1.00 – 1.01) | 0.311 |
| **Logit Model** | | |
| **(Intercept)** | 0.04<br>(0.01 – 0.24) | **0.001** |
| Screentime | 1.85<br>(1.18– 2.90) | **0.007** |
| Day | 1.08<br>(1.04 – 1.13) | **<0.001** |
| Group [MA] | 0.02<br>(0.00 – 0.25) | **0.003** |
| **Random Effects** | | |
| $\sigma^2$ | 0.27 | |
| $\tau_{00\ participant}$ | 0.27 | |
| ICC | 0.50 | |
| $N_{participant}$ | 80 | |
| Observations | 1154 | |
| Marginal $R^2$/Conditional $R^2$ | 0.211/ 0.607 | |

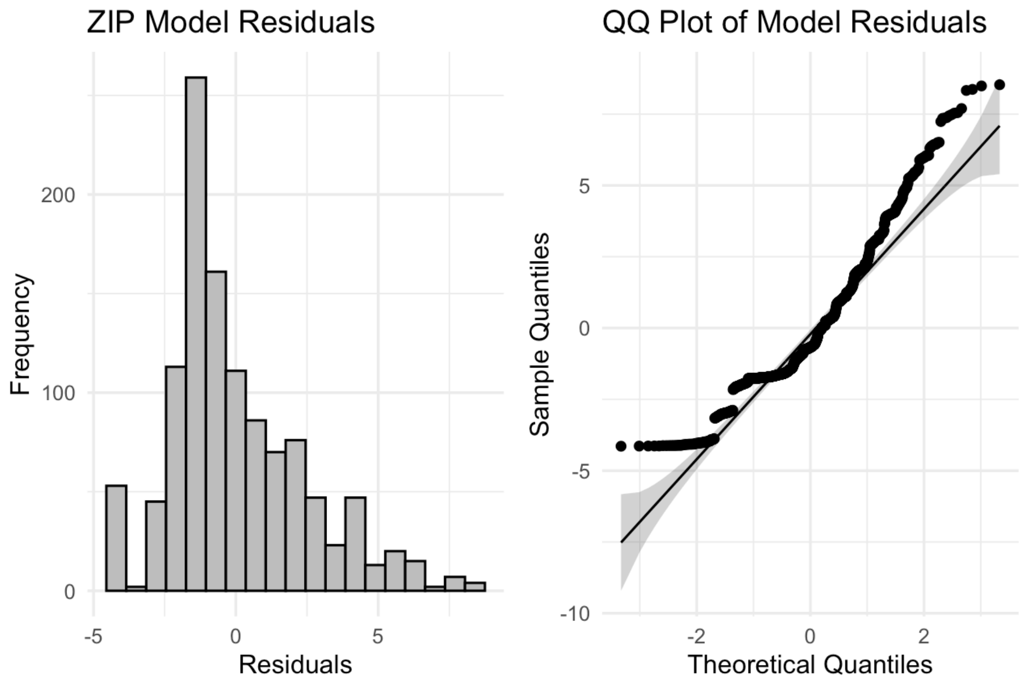
ZIP Model Residuals
Frequency
200
100
0
-5
0
5
Residuals
QQ Plot of Model Residuals
Sample Quantiles
5
0
-5
-10
-2
0
2
Theoretical Quantiles

**Table 15.** Association between real-world depression and screentime

| Predictors | Real-World Depression ~ Screentime + day + group + (1\|participant)<br>Logit: ~ screentime + day + group + (1\|participant)<br><br>*Incidence Rate Ratios* | *p* |
|---|---|---|
| **Count Model** | | |
| (Intercept) | 1.40<br>(1.04 – 1.87) | **0.024** |
| Screentime | 1.01<br>(0.96 – 1.07) | 0.625 |
| Group [MA] | 2.81<br>(2.01 – 3.92) | **< 0.001** |
| Day | 1.00<br>(0.99 – 1.01) | 0.851 |
| **Logit Model** | | |
| **(Intercept)** | 0.05<br>(0.00 – 1.53) | 0.085 |
| Screentime | 1.08<br>(0.63 – 1.88) | 0.776 |
| Day | 1.09<br>(1.03 – 1.16) | **0.006** |
| Group [MA] | 0.00<br>(0.00 – 0.15) | **0.003** |
| **Random Effects** | | |
| $\sigma^2$ | 0.28 | |
| $\tau_{00\ participant}$ | 0.31 | |
| ICC | 0.53 | |
| $N_{participant}$ | 80 | |
| Observations | 1146 | |
| Marginal $R^2$/Conditional $R^2$ | 0.301/ 0.669 | |

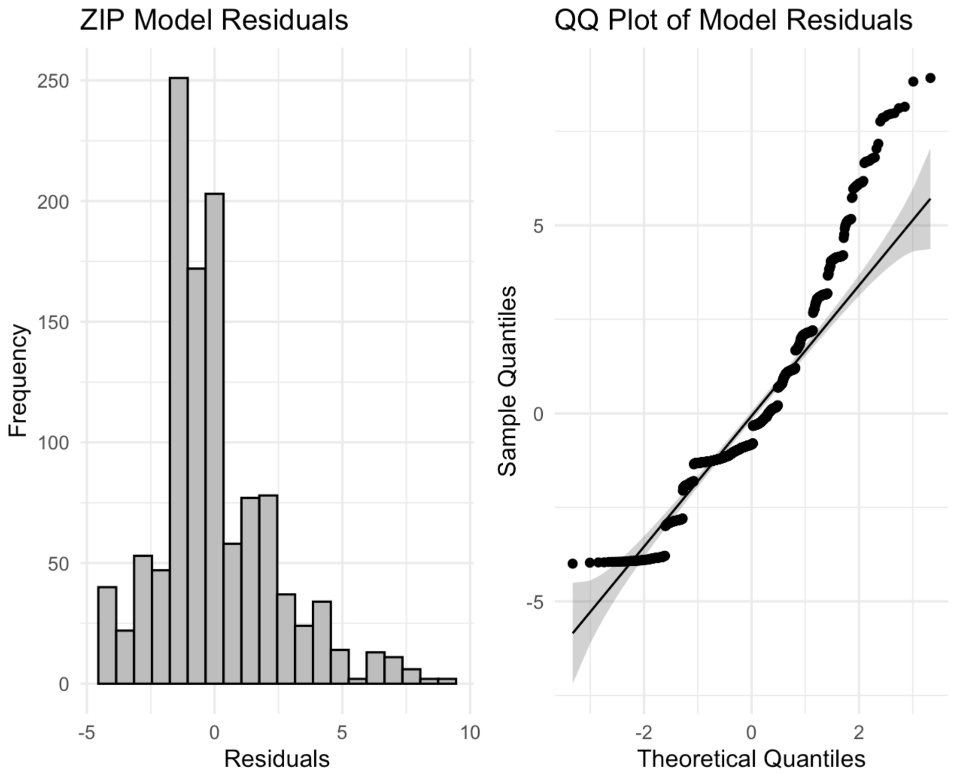
ZIP Model Residuals
Frequency
250
200
150
100
50
0
-5
0
5
10
Residuals
QQ Plot of Model Residuals
Sample Quantiles
5
0
-5
-2
0
2
Theoretical Quantiles
